# Health care utilization of fine-scale identity by descent clusters in a Los Angeles biobank

**DOI:** 10.1101/2022.07.12.22277520

**Authors:** Christa Caggiano, Arya Boudaie, Ruhollah Shemirani, Ella Petter, Alec Chiu, Ruth Johnson, Defne Ercelen, Bogdan Pasaniuc, Eimear Kenny, Jonathan Shortt, Chris Gignoux, Brunilda Balliu, Valerie Arboleda, Gillian Belbin, Noah Zaitlen

## Abstract

An individual’s disease risk is affected by the populations that they belong to, due to shared genetics and shared environment. The study of fine-scale populations in clinical care will be important for reducing health disparities and for developing personalized treatments. In this work, we developed a novel health monitoring system, which leverages biobank data and electronic medical records from over 40,000 UCLA patients. Using identity by descent (IBD), we analyzed one type of fine-scale population, an IBD cluster. In total, we identified 376 IBD clusters, including clusters characterized by the presence of many significantly understudied communities, such as Lebanese Christians, Iranian Jews, Armenians, and Gujaratis. Our analyses identified thousands of novel associations between IBD clusters and clinical diagnoses, physician offices, utilization of specific medical specialties, pathogenic allele frequencies, and changes in diagnosis frequency over time. To enhance the impact of the research and engage the broader community, we provide a web portal to query our results: www.ibd.la

## 2 Introduction

Individuals belong to many populations, and each population can have unique health risks. This can be a consequence of a population’s shared cultural or physical environment, shared genetics, or a combination of both. Understanding population-level differences in disease outcomes will be important for reducing health disparities and for developing personalized treatments [1], [2]. To this end, new large-scale biobanks tied to electronic health records (EHR) present an ideal opportunity to study the health of populations [3]–[5]. These biobanks can facilitate the study of many types of populations, especially groups of people who are genetically related. For example, recent work in biobanks has identified new genetic associations [6], examined how diseases track through families [7], and developed new polygenic risk scores for diverse ancestries [8]–[10].

Our work [11], along with others [12]–[15], has used identity-by-descent segments (IBD) to identify fine-scale populations in biobanks. Here, we use one definition of population, which are clusters of individuals who share IBD. IBD segments are stretches of DNA that are identical between individuals due to having a shared ancestor. People whose ancestors lived in the same geographic location or who were part of the same ethnolinguistic group tend to share more IBD [16]. These clusters of people may also share an environment, which can be relevant for understanding why or how patients visit the hospital. Indeed, our previous work demonstrated that groups of individuals who share elevated amounts of IBD are associated with particular diagnoses. Furthermore, IBD clusters can complement measures of self-reported race and ethnicity and offer additional information on genetic risk. IBD analysis, then, can be a powerful tool for identifying opportunities to improve health outcomes.

Here, we extend our previous work [11], and use IBD to detect fine-scale clusters and analyze their health patterns within the ATLAS Community Health Initiative [17]. ATLAS is part of the University of Los Angeles (UCLA) Health system located in west Los Angeles, which serves a diverse population of patients across the greater Los Angeles area. Los Angeles is a cosmopolitan city, characterized by many neighborhoods whose residents represent recent and past immigration into California (Fig. 1a) [18]. By creating a network of IBD sharing and applying an unsupervised machine learning algorithm, we identified 376 distinct IBD clusters in ATLAS. IBD clusters include groups defined by the presence of Iranian Jews, Armenians, Lebanese Christians, Egyptian Christians, Filipinos, Puerto Ricans, Koreans, and Ashkenazi Jews. Many of these groups have been historically understudied in biomedical research. Thus, this work represents an advance in the study of the health of minority groups in Southern California. Note that for this work, we name the IBD clusters to facilitate communication. We emphasize, however, that grouping and naming are by nature, error-prone, especially in the context of continuous human diversity [19]. Furthermore, not all individuals within a cluster will share the same attributes, including self-identified race, ethnicity, language, or religion. As such, IBD clusters offer one lens into studying health outcomes that can be studied alongside socially determined concepts of race and ethnicity to better understand structural determinants of health [20], [21].

**Figure 1:**
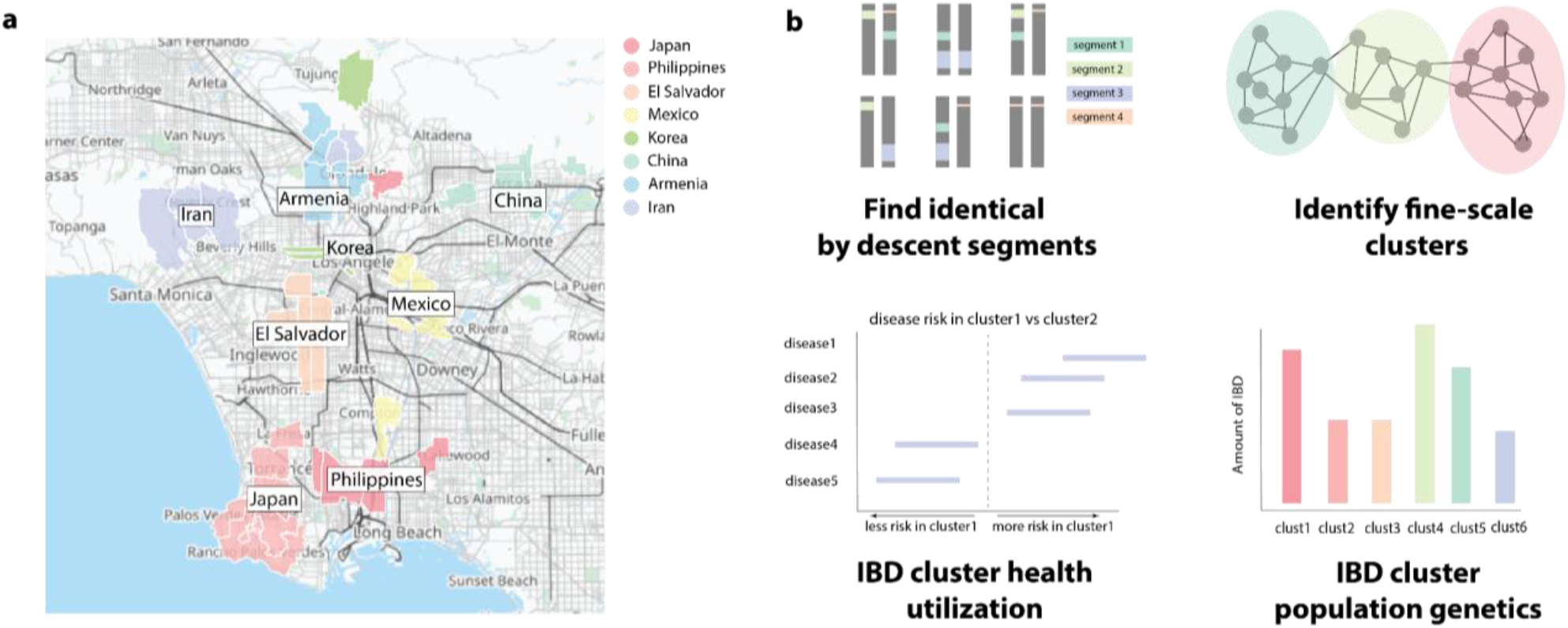
Foreign-born residents in Los Angeles and an overview of the fine-scale cluster detection approach. (a) For eight relevant countries, the top five zip codes in LA county with the most individuals born in that country according to 2019 US Census Data. (b) A schematic of IBD calling and cluster annotation. We first pre-process the data and infer IBD segments for all biobank participants and reference samples (top left). We then identify fine-scale clusters using Louvain clustering (top right) and we use EHR data associated with the individuals in each IBD cluster to explore patterns of enrichment for cluster-specific health utilization (bottom left). Finally, we measure patterns of genetic relatedness both within and between identified clusters (bottom left).

We identified thousands of novel cluster-specific health associations, especially in the Iranian, Armenian, and South Asian IBD clusters. There were widespread differences in the rates and patterns of interaction with the healthcare system, including the observation that non-European groups were less likely to visit a doctor that had a primary care specialization. Differences in diagnoses between clusters included elevated rates of heart transplants in the Armenian cluster, eating disorders in the Ashkenazi Jewish cluster, and non-toxic multinodular goiters in both Iranian clusters. We also examined the frequencies of pathogenic alleles within IBD clusters and noted several instances of cluster-specific enrichment, including a high carrier rate for familial Mediterranean fever in Armenians, and a rare allele for GNE myopathy in Iranian Jews. Finally, we analyzed the genetic properties of IBD clusters. Several clusters had high within-cluster IBD sharing, indicating elevated relatedness. In particular, the Iranian Jewish cluster shared an average of 57.43 cM of IBD, which is much higher than estimates for other well-known founder groups, such as the Ashkenazi Jewish (26.08 cM) and Puerto Rican (23.06cM) clusters. To facilitate the use of the large set of novel associations, we developed a web framework allowing interactive access to the results presented here. Overall, this work progresses understanding of health in understudied communities, which can be useful in the development of precision medicine and can provide the foundation for further study into structural health inequities that may exist in Los Angeles.

## 3 Results

### 3.1 ATLAS

The UCLA ATLAS Community Health Initiative aims to create a genomic resource to enable translational and precision medicine (see ref [17] and [6] for more details). In ATLAS, genotyping data is tied to de-identified EHRs as part of the UCLA Health IT Discovery Data Repository & Dashboard (DDR) [22]. As of 2021, there were approximately 40,000 participants with full genotyping and DDR data available [23]. Recruitment is ongoing. The DDR includes records from the large UCLA Health system, which includes two main hospital campuses, the Ronald Reagan UCLA Medical Center located in Westwood, and the UCLA Santa Monica Medical center UCLA Health also has more than 200 clinics throughout the Los Angeles area with thousands of affiliated physicians. The DDR includes demographics, diagnoses, records of encounters containing information on the provider and office of a visit, prescription medication orders, and procedures. ATLAS comprises a diverse set of individuals, both genetically, and in terms of demographic characteristics reported in the EHR (Fig. S1). A complete description of the ATLAS project and data is available in [17].

### 3.2 Identifying fine-scale ancestry clusters

#### 3.2.1 Use of genetic ancestry in determining clusters

To identify fine-scale clusters that are relevant for studying health outcomes, we focused on inferring relationships via IBD segments (Fig. 1b). Our previous work showed that studying IBD clusters offer distinct advantages over identifying patient clusters through self-reported or EHR-reported measures alone. This was especially true for non-European ancestry populations [11]. We find similar properties here. 11.6% of patients in ATLAS do not have a race/ethnicity specified. Of those that do have a race/ethnicity specified, 14.7% have the designation of “Other Race,” which is the second most common race/ethnicity designation after “White” in ATLAS. This category can encompass many non-European groups, including those with Middle Eastern ancestry, who are a key demographic group in Los Angeles. Thus, using race/ethnicity categories alone limits our ability to study health outcomes that may exist in non-European groups.

While other characteristics, such as preferred language, could be used to study determinants of health, many patients do not have this information recorded in the EHR. For example, only 5.2% of patients prefer a language other than English, which would limit the power to detect disease associations. Additionally, the reasons an individual may have language, or other similar demographic information, included in their EHR record is complex, non-random, and not guaranteed to be accurate. Therefore, for this study we focus on groups identified via genetic ancestry. Importantly, genetic ancestry is a distinct concept from race, which is a social construct [19], [24]. The study of both can offer valuable and complementary perspectives toward the goal of understanding the reasons a group visits a hospital [25]. Future studies could focus on further expanding self-identification surveys in the context of EHRs, which could work in tandem with genetic ancestry clusters to identify health disparities.

#### 3.2.2 Identity by descent clustering

To define our IBD clusters, we called pairwise IBD between all ATLAS participants and reference individuals sourced from the 1000 Genomes Project [26], the Simons Genome Diversity Project [27], and the Human Genome Diversity Project [28]. IBD segments were estimated using iLASH [29], identifying in total, over 95 million shared IBD segments. All participants in the biobank had at least one IBD segment detected, with the mean amount of IBD sharing being 14.80cM (IQR: 3.84-21.57cM). This was similar to the amount of IBD sharing in our previous work using the BioMe biobank from the Mount Sinai Health System, which found a mean 19.10cM (IQR: 5.61-27.52cM) [11]. In ATLAS, total IBD ranged from 3cM to more than 100cM, with about 30% being less than 4cM, indicating both substantial ancient and recent genetic relatedness (Fig. S2a). Most pairs only shared 1 segment of IBD (Fig. S2b).

To infer clusters, we followed the approach of Dai et al. and used the Louvain method for cluster detection [30]. This method finds structure in large networks and has been shown to work well on genetic data [13]. We applied this algorithm to an undirected network constructed from IBD sharing, where each node represented an individual, and edge weights were defined as the genome-wide sum of IBD sharing between the nodes [31]. An advantage of the Louvain algorithm is that it can be run iteratively, meaning that an initial run over the entirety of the graph can be used to define broad substructure, which can be further resolved into more fine-scale clusters upon subsequent iterations. We detected 376 IBD clusters, each of which was given a provisional identifier corresponding to the cluster from each of the three iterations of Louvain clustering (e.g., “UCLA_1_0_2”). The mean number of patients per cluster was 106.85±280.05, which was about 0.3% of ATLAS. There was substantial variation in the size of the estimated clusters, ranging from a minimum of 2 individuals to maximum 2030 individuals, which was 5.6×10^−5^ % to 5% of ATLAS. Overall, these clusters represented substantial genetic diversity, with clusters characterized by a mixture of ancestries from the Americas, West Asia, Europe, Africa, East Asia, and South Asia (Fig. 2a, Fig. S3). Differences in cluster size, historic population size, and complex patterns of genetic relatedness resulted in differential cluster densities. Some clusters are densely connected, with nearly every individual sharing IBD with every other individual in the group (Fig. S4a), while others share few connections (Fig. S4b), or a moderate amount (Fig. S4c).

**Figure 2:**
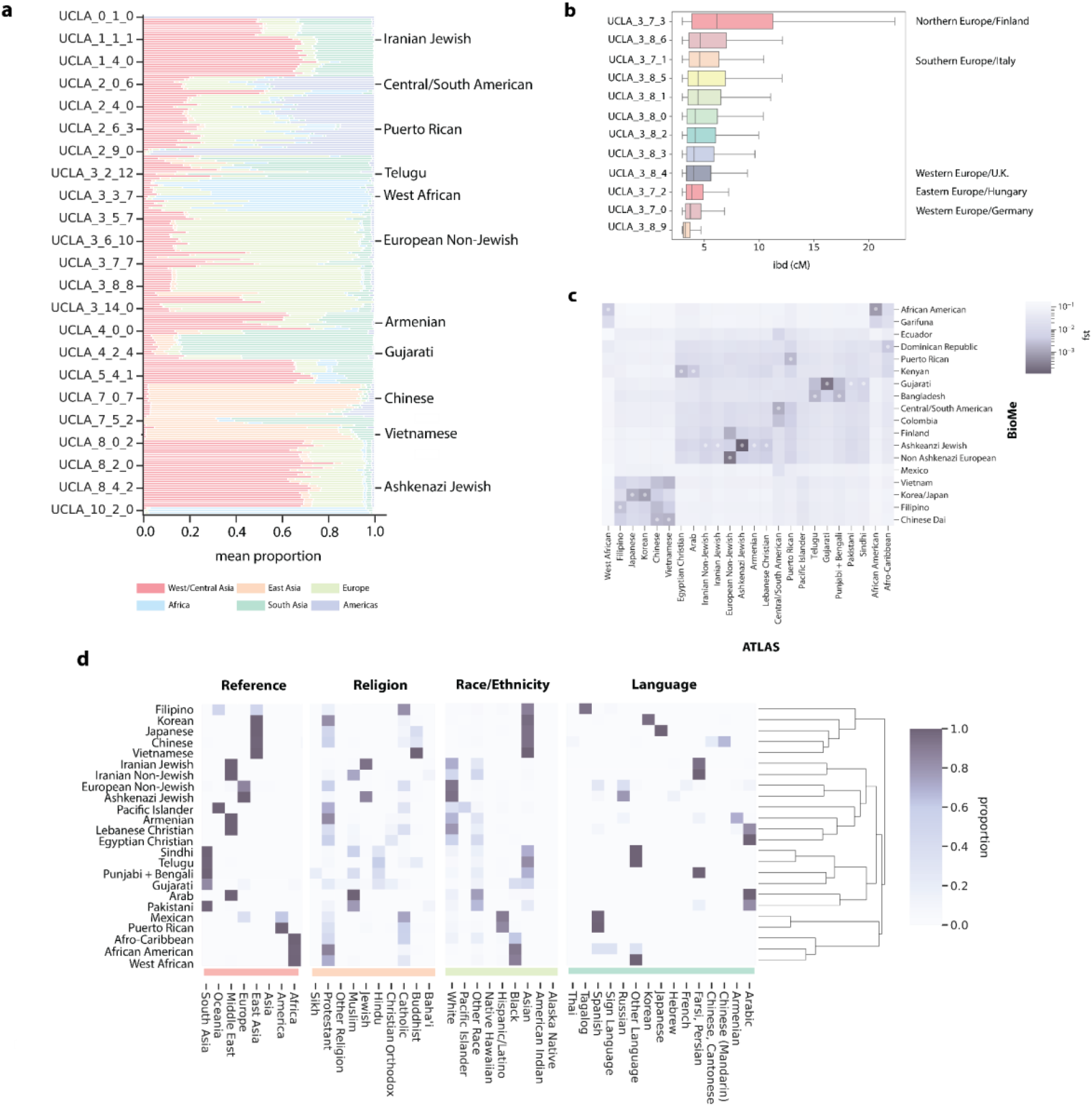
Genetic and demographic properties of clusters. **(a)** The mean admixture fractions [33] for each of the UCLA ATLAS IBD clusters when K=6. Each line corresponds to the components for one ATLAS IBD cluster. The components are inferred to correspond to genetic ancestry from the Middle East (red), East Asia (orange), Europe (green), South/Central Asia (teal), Africa (blue), and the Americas (purple). The left column indicates the number of the IBD cluster from the Louvain algorithm. The right column gives examples of names that were given to the largest Louvain IBD clusters. **(b)** The distribution of IBD within subclusters that were merged to make one cluster of predominantly European ancestry. The names on the left indicate the number of the IBD cluster from the Louvain algorithm, and the name on the right indicate potential names from F_ST_ analysis with the UKBB (Fig. S4a). **(c)** The Hudson’s F_ST_ value between IBD clusters identified in BioMe at Mount Sinai and ATLAS IBD clusters demonstrating the relationship between ATLAS and populations outside of Los Angeles. The darker the color, the smaller the F_ST_ value. The smallest F_ST_ value for each of the ATLAS clusters is indicated by a white dot. **(d)** For each of the largest clusters, (from left to right) the proportion of reference data by continent in each cluster, the proportion of the cluster that indicated they prefer a specific religion, the proportion of the cluster marked in the EHR as each race/ethnicity category, and the proportion of each language preferred by the cluster.

To further refine our clusters, we used the approach of Dai et al. and merged subclusters with low F_ST_ (Hudson’s *F*_*st*_ *<* 0.001) between them [13]. This facilitated identifying clusters that were differentiated enough to represent the underlying diversity of ATLAS, but sufficiently large to allow for powered statistical analysis. This stopping point, however, is arbitrary, and finer-scale clusters might be interesting for future population or medical genetics research. For example, the subclusters merged together to make the predominantly European ancestry cluster have differing distributions of IBD. F_ST_ to an external dataset of UK BioBank (UKBB) participants born outside the UK suggest that these subclusters likely represent individuals from different geographic regions, such as Northern, Southern, and Eastern Europeans (Fig 2b). In total after F_ST_ merging, we identified 24 clusters of diverse ancestry with at least 30 ATLAS participants, representing 97.8% of ATLAS. For downstream analyses, we focus on these 24 clusters.

#### 3.2.3 Annotating cluster identity

We next assigned names to each cluster. The ATLAS biobank does not contain the true country of origin of participants, which was used in our previous studies to annotate cluster identity [11]. Instead, we annotated IBD clusters by using reference data in the clustering algorithm. We also used additional summary statistic reference data to compute Hudson’s F_ST_ between ATLAS IBD clusters and external populations (Fig. 2c, Fig. S4), including IBD clusters identified in the BioMe biobank. This enabled additional refinement of cluster names. Whenever possible, we named the IBD clusters according to previously published naming conventions.

Some clusters did not contain any reference data, or the reference data did not capture important aspects of the cluster. For example, there was no Ashkenazi Jewish reference data, only reference data labeled by European countries. To address this problem, we used the de-identified EHR demographic information to refine and confirm cluster annotations. These included EHR reported race and ethnicity, preferred language, and preferred religion (Fig. 2d). We emphasize that race, ethnicity, and religion are not determined by IBD segments, but represent sociocultural characteristics that may be related to cluster identity. We chose to use religion in the name of groups when it was relevant to identifying a historically persecuted group (i.e., “Lebanese Christian” instead of just “Lebanese”). These groups often have distinct histories and cultural practices, which can affect demography, environment, and disease risk. For example, it is well known that Ashkenazi Jews have distinct genetic risks relative to other Europeans [32]. Thus, including religion in this study may offer opportunities to improve the health of understudied ethnoreligious groups.

Most ATLAS patients are non-Hispanic, have no religious preference, and indicated that they prefer to speak English. We, therefore, labeled clusters using individuals who preferred a different language or religion or were identified as Hispanic in the EHR (note that the actual number of English speakers may be lower, as some patients may not, for societal or practical reasons, have this information included in their medical records). Importantly, the label given to a cluster serves as a broad interpretation of the cluster’s demographic ties and does not necessarily reflect the self-identity of members, see Discussion for further details. Furthermore, the clusters discussed here are specific to Los Angeles, especially those who visit UCLA Health, and may not be representative of the global population.

We identified many fine-scale clusters that reflect the known demography of Los Angeles. For example, we identified an Iranian Jewish (n=264) and an Iranian Non-Jewish IBD cluster (n=350), which are groups that predominantly immigrated to Los Angeles after the 1979 Iranian Revolution [34]. We also identified a large Armenian cluster (n=491), consistent with Los Angeles having the largest population of diaspora Armenians in the US [35]. Furthermore, we identified several Asian clusters, including Chinese (n=1547), Filipino (n=796), Vietnamese (n=269), Japanese (n=596), and Korean (n=546) clusters. While Spanish-speaking reference data was lacking, we still identified IBD clusters characterized by the presence of Puerto Rican (n=288) and Colombian (n=49) reference samples. One Spanish-speaking cluster, which we call “Central/South American,” was large (n=6075), and contained reference data from a variety of groups, including Mexicans, and Peruvians. Subgroups of this cluster showed complex fine-scale structure (Fig. S4E), suggesting that in the future, it could be resolved into further subclusters.

Other notable IBD clusters were named Ashkenazi Jewish (n=5309), Lebanese Christian (n=219), Egyptian Christian (n=92), and Telugu (n=276). We identified three Black/African American clusters. One cluster, termed “African American”, was the largest (n=1877). It contained individuals with significant admixture components of both European and African ancestry and included the 1000G African American reference samples. The second IBD cluster, “Afro-Caribbean” (n=39), had low F_ST_ to the Dominican cluster in BioMe (Fig. 2C) and other Caribbean populations in an external dataset of foreign-born participants in the UKBB (Fig. S4F). The third IBD cluster, which we named “West African” (n=281), had a large African admixture component and clustered with Yoruba reference samples, which suggested that this cluster represented individuals with more recent African ancestry than the other Black/African American IBD clusters. Further details about the cluster characteristics and associated reference data can be found in Supplementary Data 1.

### 3.3 Health system utilization of fine-scale IBD clusters

#### 3.3.1 Phecodes

Using de-identified EHR data, we sought to understand how different IBD clusters accessed the hospital system and what conditions they presented with. This may be of interest to health care practitioners seeking to improve care for diverse groups and provides an opportunity to understand health disparities that may exist within Los Angeles.

To assess whether an IBD cluster received a diagnosis at a higher or lower rate than others, we used a logistic regression model, regressing whether an individual received a particular diagnosis against the individual’s IBD cluster status. Phecodes based on ICD10 codes were used in place of diagnoses, as they have been shown to be good proxies for disease phenotypes [36]. To account for differences in diagnosis frequencies between medical contexts, we separately assessed the code assignments both for outpatient encounters and for emergency room (E.R.) visits. Age, sex, and BMI were controlled for in both analyses. We note that there was substantial variation between the clusters for all these factors (Fig. S5), further highlighting the structural and environmental factors shape health amongst ATLAS patients.

We began by comparing phecode assignments in Ashkenazi Jews to all biobank participants in an outpatient setting. Ashkenazi Jews represent one of the largest IBD clusters identified (n=5309) and are relatively well studied, which enables validation. We tested n=1131 phecodes assigned to at least 30 patients in outpatient encounters. 236 were significantly associated with Ashkenazi Jew IBD cluster status at a false discovery rate of 5% (Fig. 3a). As reported in [11], [37], [38], we find that Ashkenazi Jews are more likely to be assigned phecodes for ulcerative colitis (log odds ratio: 0.81±0.2, p-value: 6.61e-15), chronic ulcerative colitis (log odds ratio: 0.74±0.23, p-value: 6.88e-10) and regional enteritis (log odds ratio: 1.08±0.20, p-value: 4.23e-27). We additionally observed strong associations for several mental health disorders, including obsessive-compulsive disorder (log odds ratio: 1.13±0.33, p-value: 2.49e-11), eating disorder (log odds ratio: 1.22±0.32, p-value: 1.02e-13), anxiety disorder (log odds ratio: 0.53±0.07, p-value: 8.75e-55), and major depressive disorder (log odds ratio: 0.48±0.1, p-value: 9.02e-23).

**Figure 3:**
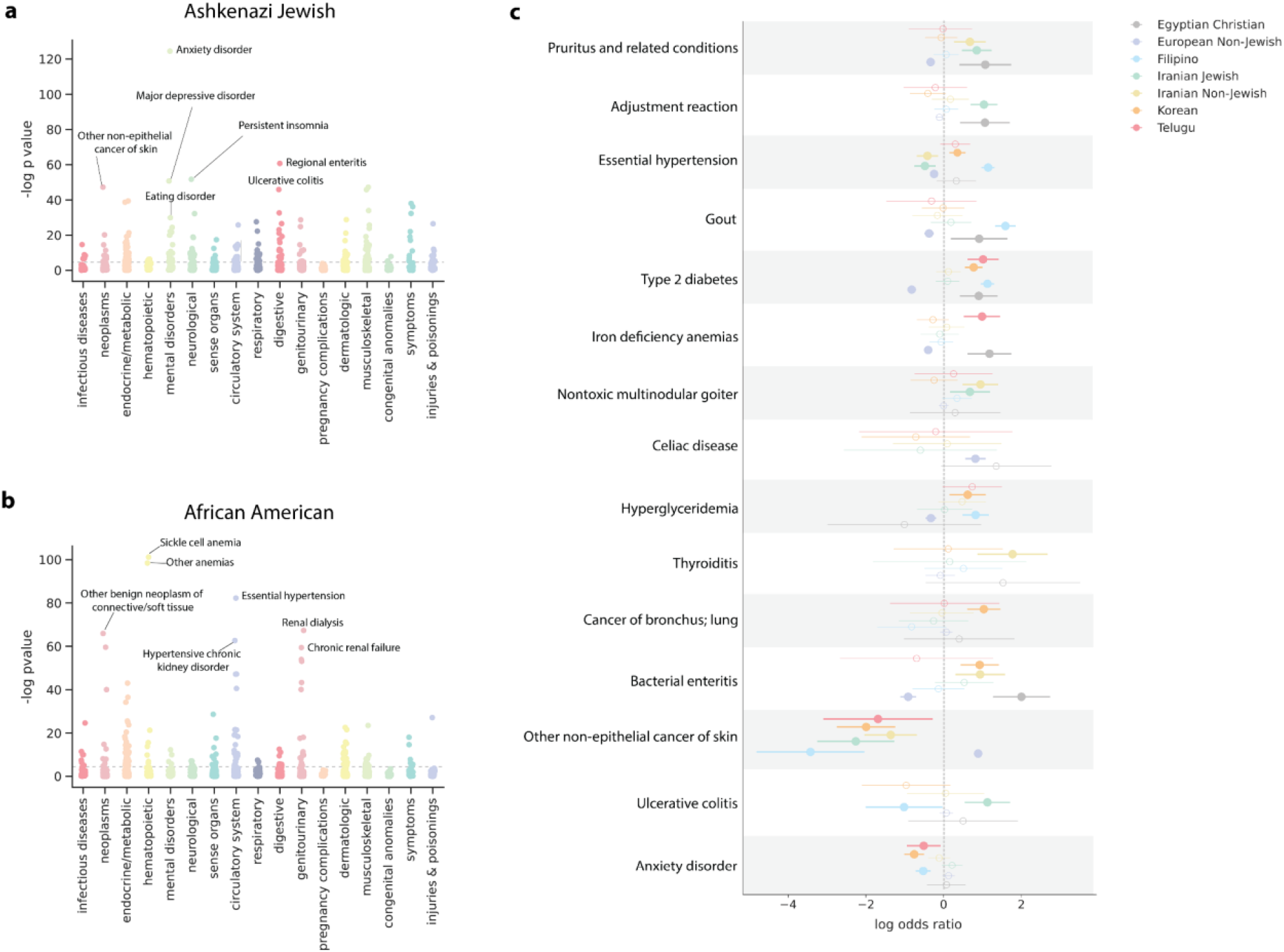
Phecode associations for selected IBD clusters. **(a)** Manhattan plot showing associations between IBD cluster status and being assigned a phecode (n=1131) relative to the remaining biobank participants. Results are shown for **(a)** the Ashkenazi Jewish IBD cluster or **(b)** the African American IBD cluster status. Phecodes are grouped by phenotypic category. The top significant (FDR 5%) for each IBD cluster are labeled and the bonferroni significance is indicated by a grey dotted line. **(c)** For 7 IBD clusters, including 6 understudied communities, the log odds ratio of the association between IBD cluster status and being assigned a phecode. A log-odds ratio greater than zero means that the phecode is more likely to be given to patients of that IBD cluster. The dot indicates the log odds ratio for the cluster and the bars indicate the 95% confidence interval. A solid dot (versus an open dot) indicates that the association between IBD cluster and phecode is significant at FDR 5%.

All these associations remained significant at FDR 5% when restricting the analysis to only compare the Ashkenazi Jewish and European Non-Jewish IBD clusters. The mental health-related enrichments were especially interesting given that in E.R. visits, Ashkenazi Jewish IBD cluster status was significantly associated with major depression as the primary reason for visit (log odds ratio: 0.83±0.55, p-value: 0.0034) (Fig. S7a). While these results are consistent with previous reports of mental illness in Jewish communities of European ancestry [39]–[41], we emphasize that this association does not indicate a causal relationship between Jewish IBD cluster status and these disorders. Instead, this association could reflect a shared environment, such as social pressure, including response to anti-Semitism as suggested in [42], socioeconomic status, or group norms around seeking psychiatric care.

We confirmed several other established disease-ancestry associations. For example, we observed that compared to the whole biobank, individuals in the African American IBD cluster (n=1877) were more likely to be assigned phecodes for sickle cell anemia (log odds ratio: 3.92±0.55, p-value: 1.18e-44) and less likely to receive phecodes for skin cancer (log odds ratio: -3.57±1.96, p-value: 3.61e-04) (Fig. 3b). Both these observations are consistent with previous studies [43], [44]. Notably, this IBD cluster is also more likely to have pathogenic alleles in the HBB gene, which is responsible for sickle cell disease [45] (see Section 3.4 for further discussion). Furthermore, as with previous reports [46], we observed an increased number of diagnoses relating to Viral Hepatitis B in Asian IBD clusters. However, the effect size significantly differed (Mantel-Haenszel chi-square test, p-value=5.21e-06) between the Chinese (n=1547) (log odds ratio: 2.95±0.25, p-value: 1.66e-120), Korean (n=546) (log odds ratio: 2.16±0.37, p-value: 4.16e-31), and Filipino (n=767) (log odds ratio: 1.14±0.48, p-value: 4.00e-06) IBD clusters, indicating the utility of fine-scale information.

Next, we focused on identifying disease associations in understudied communities (Fig. 3c). The Iranian Jewish (n=264) and Iranian Non-Jewish (n=351) IBD clusters shared several associations in outpatient diagnoses. Both communities were less likely to be diagnosed with certain types of skin cancer and more likely to be assigned codes relating to goiters. However, the phecode with the smallest p-value for each cluster-non-toxic multinodular goiter in Iranian Non-Jewish (log odds ratio: 0.94±0.46, p-value: 4.83e-05) and adjustment disorder in Iranian Jews (log odds ratio: 1.06±0.35, p-value: 2.04e-09) were not the same. This difference emphasizes that fine-scale information is useful when studying health outcomes in Los Angeles. Other notable associations we identified included an enrichment of phecodes relating to bacterial enteritis in the Egyptian Christian IBD cluster (n=92) (log odds ratio: 2.00±0.73, p-value: 9.18E-08) and phecodes relating to bronchus cancer in the Korean cluster (n=546) (log odds ratio: 1.03±0.43, p-value: 2.03e-06).

In emergency room diagnoses, we observed a particularly high number of severe diagnoses associated with the African American cluster status (Fig. S7b), including phecodes relating to pulmonary heart disease (log odds ratio: 1.19±0.37, p-value: 4.21e-10), chronic renal failure (log odds ratio: 1.15±0.4, p-value: 2.50e-08), and cardiomyopathies (log odds ratio: 1.14±0.44, p-value: 4.39e-07). All of these conditions are well documented to be at a higher prevalence in the general African American population in the United States [47]–[49]. The Central/South American IBD cluster was more likely to visit the emergency room for reasons relating to liver transplants (log odds ratio: 1.51±0.3, p-value: 1.13e-23). Interestingly, the Chinese IBD cluster, who despite the relatively large cluster size, had fewer emergency room visits (log odds ratio: -0.29±0.068, p-value: 1.97e-05) (Supplementary Table 2) than other clusters, and no significant phecode associations against the entire biobank. This does not mean that Chinese individuals are less likely to have health emergencies, but rather, could reflect other structural factors, or that this cluster visits other emergency rooms that are not in the UCLA Health System.

We noted that phecode associations depended strongly on the set of comparator clusters. While it is straightforward to consider the phecodes assigned to an IBD cluster relative to the entire biobank, researchers might also be interested in enrichments between clusters that are closely related. However, “closely related” depends on a complex interplay of culture, environment, and perception. For example, Armenia is in Western Asia and has a strong historical relationship with Iran [50]. Armenia, however, also has close ties with Europe [51], and 68% of the Armenian cluster is designated as “white” in the UCLA EHR. When comparing the Armenian IBD cluster (n=491) against the entire biobank, they were more likely to be assigned codes relating to heart transplant (log odds ratio: 1.39±0.5, p-value: 4.72e-08), which is consistent with previous reports of Armenian ancestry being a risk factor for cardiovascular disease [52] (Fig. 4a).

**Figure 4:**
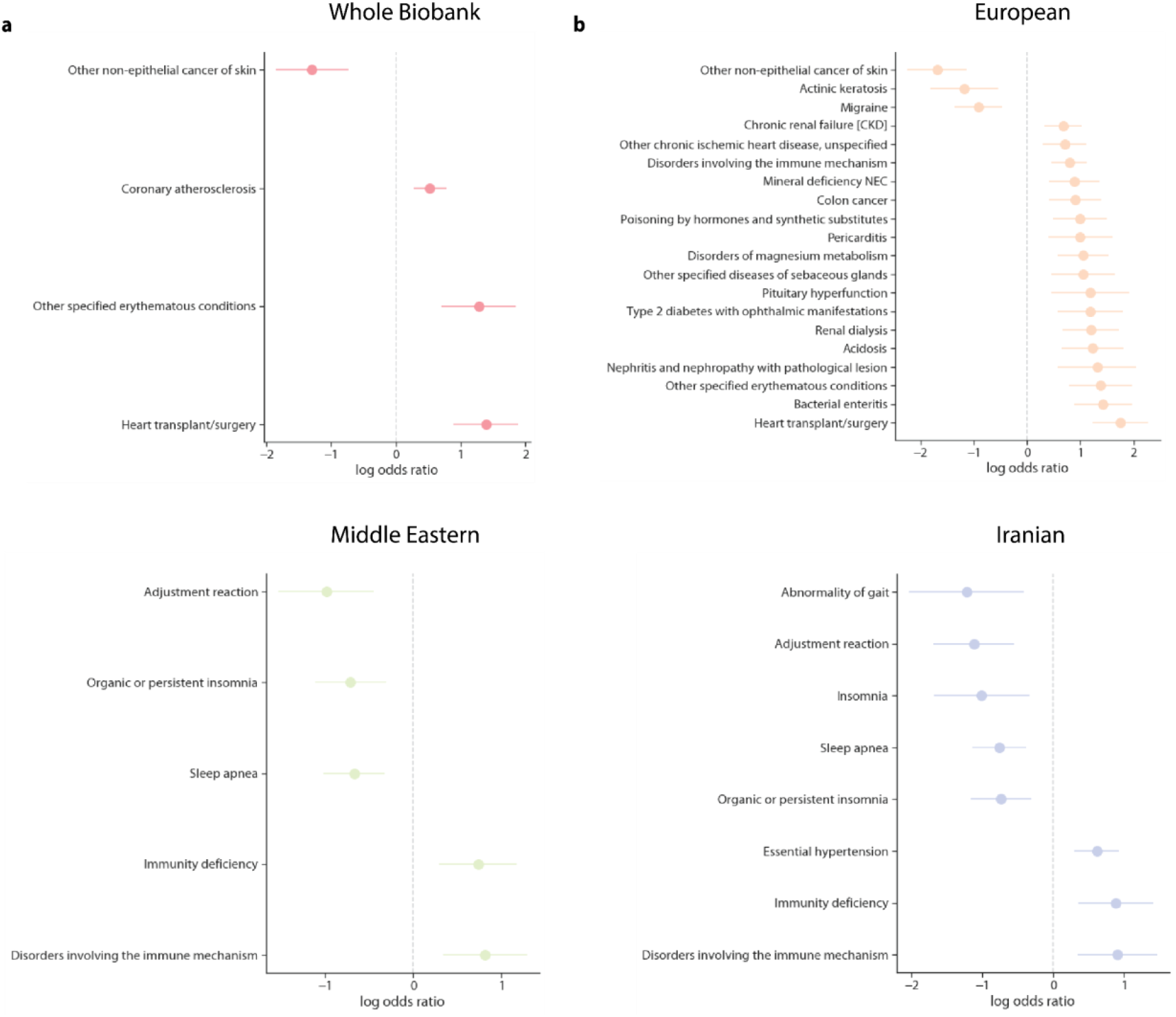
Phecodes associated with the Armenian IBD cluster compared to different populations. The log odds ratio of a given phecode assignment is associated with membership in the Armenian cluster versus membership in **(a)** the remaining biobank at large, **(b)** the European Non-Jewish clusters **(c)** the Middle Eastern clusters (both Iranian IBD clusters, both Christian Arab IBD clusters, and the Arab IBD cluster) and **(d)** the Iranian IBD clusters only. A positive log odds ratio means that the phecode is more likely to be given to patients of that cluster. We tested only phecodes with at least 30 patients per code and corrected for age, sex, and BMI in these analyses. Phecodes significant at FDR 5% are shown and if there are more than 30 significant associations, we plot only the top 20.

When comparing the Armenian IBD cluster against the European Non-Jewish IBD cluster (n=17017) (Fig. 4b), these disorders were still significant, but many new enrichments surfaced. When comparing the Armenian cluster to Middle Eastern IBD clusters (Iranian Non-Jewish and Jewish, Egyptian and Lebanese Christians, and Arabs) (n=960) (Fig. 4c) or only the Iranian IBD clusters (n=614) (Iranian Non-Jewish and Iranian Jews) (Fig. 4d), we found that many phecodes that were enriched against the European IBD cluster were no longer significant. New phecodes were also identified, such as disorders of the immune mechanism (log odds ratio: 0.9±0.56, p-value: 1.49e-03). This may be due to reduced power but could also be a consequence of the shared genetics between Middle Easterners and Armenians, shared diaspora culture, or both. In sum, this example illustrates the importance of holistically evaluating health disparities, as they are likely determined by a multitude of structural forces.

#### 3.3.2 Department and Office Utilization

While phecodes offer a granular look at what brings clusters to the UCLA Health system, we also sought to evaluate how IBD clusters access health care on a larger scale. To do this, we assessed whether an IBD cluster was more or less likely to visit a particular specialty using a logistic regression model, regressing IBD cluster status against whether that individual had ever visited a doctor with a given specialty in an outpatient setting.

We found that many non-European IBD clusters were significantly less likely to visit a routine care provider. For example, compared to remaining biobank participants, patients in the Central/South American IBD cluster were less likely to visit a physician with an OBGYN (log odds ratio: -0.14±0.08, p-value: 8.54e-04), internal medicine (log odds ratio: -0.25±0.06, p-value: 2.28e-15), family medicine (log odds ratio: -0.13±0.07, p-value: 3.18e-04), or primary care specialty (log odds ratio: -0.3±0.07, p-value: 2.36e-15). Likewise, belonging to the African American IBD cluster was associated with being less likely to see a primary care physician (log odds ratio: -0.27±0.13, p-value: 3.93e-05), although patients in this cluster were more likely to see a family medicine physician (log odds ratio: 0.24±0.11, p-value: 1.77e-05) than others in ATLAS. Overall, individuals who belong to the European Non-Jewish IBD clusters were significantly more likely to visit a primary care physician (log odds ratio: 0.28±0.05, p-value:6.344e-30) (Fig. 5a).

**Figure 5:**
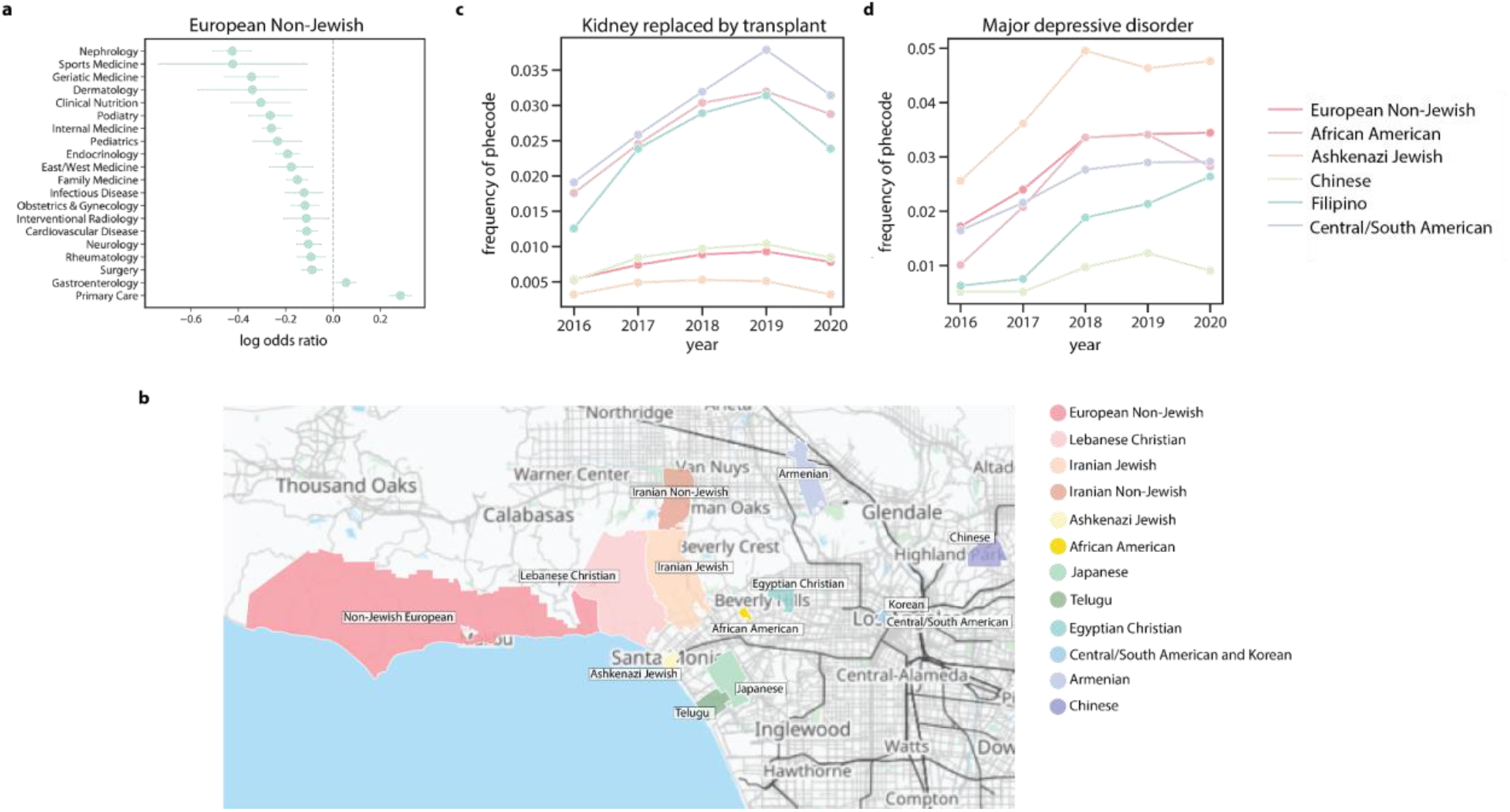
Fine-scale hospital utilization in ATLAS. **(a)** The log odds ratio of the European Non-Jewish IBD cluster having a visit to a particular specialty, assessed against all other biobank participants. **(b)** The UCLA health system office zip code which is most associated with selected IBD clusters. The color indicates what zip code is significantly associated with that cluster. For 6 clusters, the proportion of that IBD cluster that visited the UCLA Health system each year in an outpatient setting receiving **(c)** kidney replaced by transplant, and **(d)** major depressive disorder.

Beyond specialty, we also analyzed which offices within the UCLA Health System each cluster visited. The UCLA health system is large, with a network of over 2,000 physicians serving the Greater Los Angeles area [53]. Knowledge of which clusters attend specific practices might be useful for healthcare practitioners looking to better understand the populations that they serve. Furthermore, we hypothesized that patients would prefer to see doctors close to their homes, which could aid in understanding patterns of health system utilization across Los Angeles and in examining disparities in travel time to the main UCLA Hospitals.

To do this, we used a logistic regression model to test whether a patient had an encounter in a particular zip code, regressing against IBD cluster status and controlling for BMI, age, and sex. Through this analysis, we recapitulate many known geographies (Fig. 5b). For example, individuals in the Chinese IBD cluster were more likely to visit a UCLA office located in the zip code 91030 (log odds ratio: 4.22±0.6, p-value: 1.42e-43), which is situated between the cities of Pasadena and Alhambra, two major Chinese neighborhoods in California [18]. Likewise, individuals in the Korean IBD cluster were most likely to visit offices in Downtown LA adjacent to the center of historic Koreatown (log odds ratio: 0.58±0.39, p-value: 3.84e-03) [54, p.]. We also identified associations that might be less intuitive. For example, the African American IBD cluster was most likely to visit offices in the zip code 90067 (log odds ratio: 1.2±0.21, p-value: 2.52e-29), which contains a UCLA Health office located in the Century City shopping mall. This zip code has very few Black residents, (less than 2% in the 2020 census [55]), so this association might represent an instance of patients traveling to access care.

By examining the subclusters produced by the Louvain algorithm, we can potentially learn about where highly specific groups live in LA. For example, the Central/South American cluster is comprised of 5 smaller clusters that were merged after F_ST_ thresholding. These clusters could potentially represent distinct groups, such as Mexicans, El Salvadorians, and Guatemalans that may be able to be resolved as sample sizes grow and additional reference data becomes available. We found that each of these subclusters is strongly associated with a particular zip code in LA. (Fig. S8). Future work can focus on understanding structural and environmental forces specific to these zip codes to improve health.

#### 3.3.3 Diagnoses over time

We next examined how the different IBD clusters interact with the health system over time. We used phecodes as a proxy for understanding diagnoses and calculated the proportion of a cluster assigned a phecode each year. We then calculated the inter-year difference in the proportion of people diagnosed in 2020 and 2016. Since we were interested in phecodes that might have different trajectories between IBD clusters, we identified the phecodes that had the greatest variance in the inter-year difference between the 6 largest clusters. We plotted two typical codes with high variance. The proportion of patients assigned a phecode relating to kidney transplants (Fig. 5c) significantly differed between 2016 and 2019 for the Filipino (p=4.42e-05), Central/South American (p=1.77e-31), and African American (p=5.30e-07) IBD clusters, but not in the Ashkenazi, European or Chinese IBD clusters (p=9.50-04). Diagnoses generally increased but dropped sharply in 2020. This might be attributed to the strict shelter-in-place order in Los Angeles [56] and the subsequent decrease of procedures performed this year.

Phecodes relating to mental illness (Fig. 5d) were particularly interesting in their heterogeneity between clusters. The Ashkenazi Jewish IBD cluster had the highest proportion of patients diagnosed with major depressive disorder. By 2020, the Ashkenazi Jewish IBD cluster had five times as many diagnoses as the Chinese IBD cluster. The Chinese IBD cluster had a consistently low proportion receiving the phecode, and while most other clusters had an increasing number of diagnoses with time, the Chinese IBD cluster had a slow or even decreasing proportion. It is important to note that for any of these diagnoses, it is not necessarily true that the rates of diagnosis indicate the actual prevalence of the disorders in the cluster. Receiving a phecode in a UCLA hospital is extremely different than actually having a disease. Instead, these results indicate the complex dynamics between how clusters interact with the health system, which could be a function of doctor choice, insurance coverage, health care practitioner perceptions, or other structural forces.

#### 3.3.4 Disease alleles

The study of IBD clusters also presents an opportunity to identify genetic risk variants that could be included in screening efforts that have the goal of improving care for diverse patients. To explore this, we began by examining the minor allele frequency (MAF) of pathogenic mutations that have been previously reported to be enriched within particular groups. One example is Familial Mediterranean Fever, which is caused by mutations in the MEFV gene and occurs more frequently in people of Mediterranean descent [57]. We restricted to ClinVar pathogenic SNPs occurring in the MEFV gene and performed a Fisher’s exact test to compare the allele frequencies within a given IBD cluster relative to the MAF across all of ATLAS. We were limited to only the SNPs that occurred on the genotyping chip, however, which reduced the search space of pathogenic mutations. As such, there was only one pathogenic SNP genotyped in MEFV, (rs28940579), but it had significantly elevated allele frequencies in the Armenian (MAF: 0.042, Fisher’s exact test p-value: 1.72e-21), Lebanese Christian (MAF: 0.037, Fisher’s exact test p-value: 1.1 × 10-8), and Egyptian Christian IBD clusters (MAF: 0.033, Fisher’s exact test p-value: 0.00087) compared to the rest of the biobank (MAF: 5.5 × 10-3). This was consistent with expectation, as Armenians are known to have a particularly high burden of Familial Mediterranean Fever [58]. We also found that the SNP has a significantly higher minor allele frequency in the Ashkenazi Jewish IBD cluster (MAF: 0.029, fisher exact test p-value: 2.61e-159).

Next, we analyzed pathogenic variants in the HBB gene, which is implicated in sickle cell disease [45]. HBB is known to be associated with African ancestry [43]. The genotyping data contained 21 total ClinVar pathogenic SNPs in this region, and we identified two with significantly different MAF between clusters with African ancestry and the remaining biobank participants. Specifically, in the African American IBD cluster, we identified rs34598529 (biobank MAF: 3.023e-05, cluster MAF: 2.2 × 10-3, fisher exact test p-value: 1.52e-09) and rs34999973 (biobank MAF: 0.00, cluster MAF: 2.89e-4, fisher exact test p-value: 2.9 × 10-4). No variants were significant in the Afro-Caribbean cluster and only rs34598529 was significantly higher in the West African IBD cluster (cluster MAF: 0.014, fisher exact test p-value: 0.010), but this might be attributed to the much smaller sample size of these clusters.

These examples demonstrated that IBD clusters are able to recapitulate well-known disease loci, but also, we wanted to examine genetic risk loci that are not currently well-screened for. This could provide opportunities for expanding care. To test this, we examined all genotyped SNPs that were labeled in ClinVar as pathogenic and used a Fisher exact test to test for allele frequency differences between an IBD cluster of interest and the remaining biobank participants. One compelling result was rs28937594, which was significantly higher in Iranian Jews (biobank MAF: 5.80e-05, cluster MAF: 0.024, Fisher’s exact test p-value: 5.58e-28). Rs28937594 is in the GNE gene and implicated in hereditary inclusion-body myopathy, an ultra-rare recessive disease [59]. This SNP has been reported to be a founder mutation in Iranian Jews [60], [61]. Interestingly, the MAF in the Iranian Non-Jewish cluster for this SNP was high, but not significant (cluster MAF: 0.0017, Fisher’s exact test p-value: 0.1512). However, participants in the Iranian Non-Jewish cluster had a significantly higher MAF for rs41464951 (cluster MAF: 0.0051, biobank MAF:8.79e-05, Fisher’s exact test p-value: 5.02e-05), which has been found to be pathogenic for alpha thalassemia. This disease has been reported to be higher in Iranian and Middle Eastern populations [62], [63]. However, loci relating to GNE myopathy or alpha thalassemia are currently not regularly screened for in the UCLA Health System. This suggests, as others have found [64], that there are additional loci that might be included in genetic screening programs that could improve health outcomes for diverse populations.

### 3.4 Genetics of IBD clusters

#### 3.4.1 Within cluster IBD

We next turned to the genetics of IBD clusters. Studying the population genetics of a cluster can identify disease loci, as discussed in section 3.3.4, or present opportunities for learning about cultural or demographic forces, which can have implications for personalizing care or developing precision treatments [65]–[68].

First, we analyzed the IBD shared between members of the largest clusters, which can be informative about founder effects that occurred in the cluster’s past. To do this, we calculated the distribution of the total shared IBD between individuals in that cluster (Fig. 6a) (Supplementary Table 3). Of the clusters analyzed here, the Iranian Jewish cluster had the highest level of total IBD sharing (mean = 57.43 cM, 95% CI: [(56.80 - 58.06]). IBD segments shared between members of the Iranian Jewish cluster were on average 8.36cM in length (mean=8.36, 95% CI: [8.33, 8.4]) (Fig. S9). This is higher than other clusters that were expected to have founder effects, including the Ashkenazi Jewish (total pairwise IBD mean=26.08 cM, 95% CI: [26.07 - 26.09]) and the Puerto Rican IBD cluster (total pairwise IBD mean=23.06, 95% CI: [22.86 - 23.27]). The Iranian Non-Jewish IBD cluster also had relatively high IBD sharing (total pairwise IBD mean=15.70 cM, 95% CI: [14.54 - 16.86]), but not as high as the Iranian Jewish cluster. Other clusters with high within cluster IBD were the Lebanese and Egyptian Christian (total pairwise IBD mean=10.95 cM, 95% CI = [10.31 - 11.61] and mean=11.81, 95% CI: [8.8, 14.82]), the Gujarati (total pairwise IBD mean=15.93 cM, 95% CI: [15.55, 16.3]), and the Armenian (total pairwise IBD mean=10.63 cM, 95% CI: [10.24, 11.02]) clusters.

**Figure 6:**
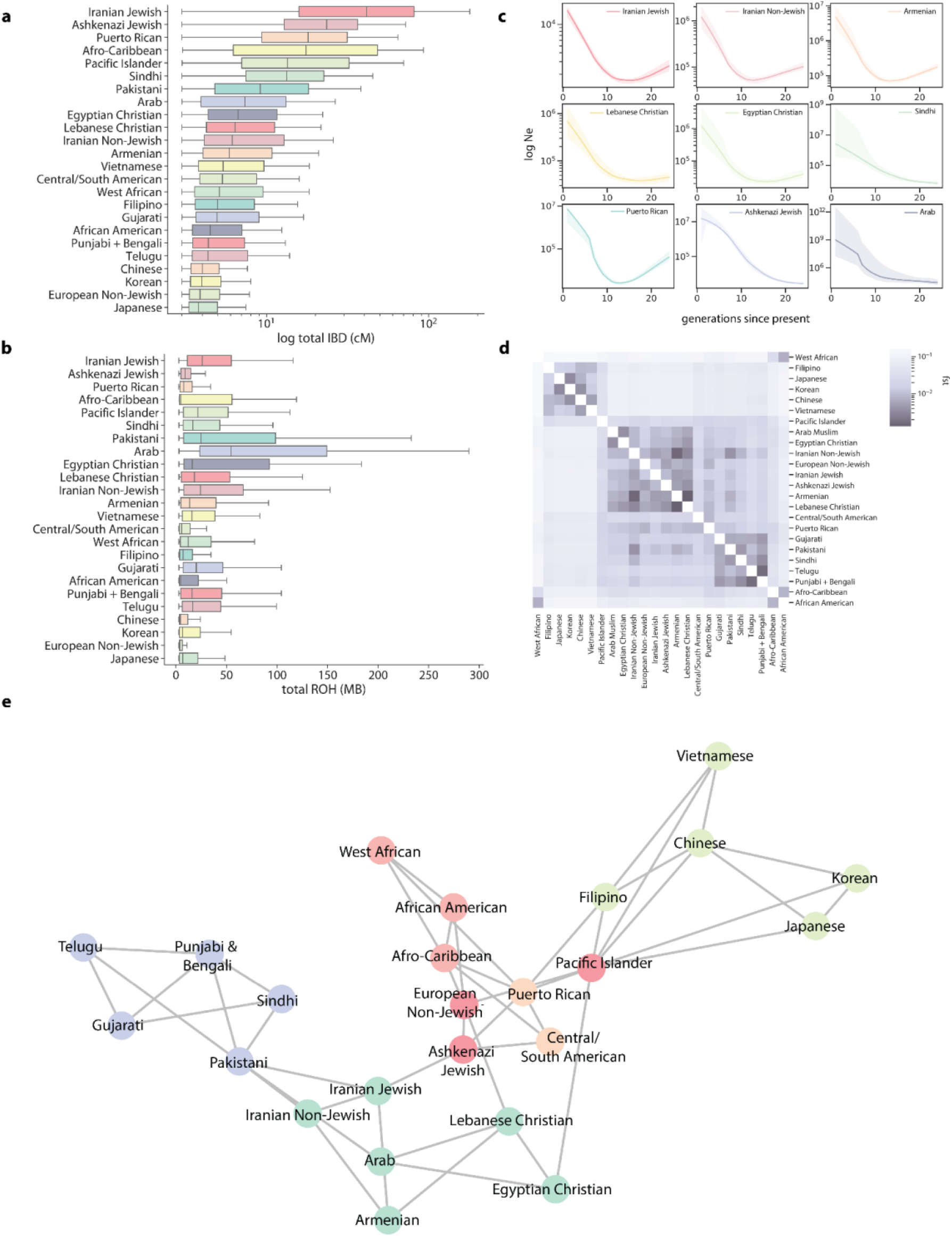
The genetic properties of the largest IBD clusters. **(a)** The distribution of total pairwise IBD (cM) shared between individuals of a given cluster. **(b)** The distribution of the total amount of ROH detected in individuals in a cluster. **(c)** IBDNe estimates of historic population size for 9 selected clusters, where the line is the estimate of the population for each generation from present, and the shaded region indicates the 95% CI of the estimate. Dips in the population size can suggest founder effects. **(d)** Pairwise Hudson’s F_ST_ estimates between UCLA ATLAS IBD clusters, where the darker color indicates lower F_ST_, suggesting less differentiation between the pair of clusters. **(e)** A network diagram of IBD sharing between clusters, where each node is a cluster, and each edge is weighted by the amount of IBD shared between the clusters. The graph was visualized using 1000 iterations of the Fruchterman-Reingold algorithm. For clarity, the 3 edges with the largest amount of IBD shared per cluster are displayed.

Additionally, we examined the distribution of runs of homozygosity (ROH) within the clusters, i.e., uninterrupted long runs of homozygous genotypes (Fig 6b). ROH have been shown to be directly implicated in risk for complex diseases [69]–[71] and relate to practices of endogamy and consanguinity [72], [73], which may be important for clinical geneticists to understand. We used PLINK to calculate the total ROH detected per individual. In doing so, we observed that 31.51% of all individuals exhibited at least one ROH, with an average total length of 18.22 MB (95% CI: [17.53, 18.90]). Upon analyzing the distributions of detected ROH within IBD clusters, we found elevated rates of ROH in several clusters. These included the Arab (mean=88.13 MB, 95% CI: [51.5, 124.75], Lebanese Christian (mean=62.44 MB, 95% CI: [36.03, 88.85]), Egyptian Christian (mean=46.07 MB, 95% CI: [32.58, 59.57]) clusters, along with several South Asian clusters. Notably, elevated cluster level IBD did not always correlate with high rates of ROH. For example, the Puerto Rican cluster had relatively low mean ROH (mean=13.27, 95% CI: [10.69, 15.85]), but high IBD. This observation may be attributed to differences in the historical demographic processes that have given rise to both measures of cryptic relatedness in the contemporary populations, for example, practices of endogamy, or the timings and magnitude of historical population bottlenecks.

To explore this further, we used IBDNe [74] to estimate cluster-specific historical effective population size and understand the population histories of clusters exhibiting high levels of IBD (Fig. 6c). This can further elucidate founder effects in groups that are not typically studied in population genetics, which can have the consequence of extending genetics care to a more diverse set of patients. Consistent with previous reports [75], we observed a large bottleneck in the Puerto Rican IBD cluster, with a minimum population size occurring around 15 generations ago. We also observed historic population size reduction in several other clusters, especially in the Iranian, Armenian, and Egyptian Christian clusters. The timing of the bottleneck in all these clusters is similar and might suggest that common historical events reduced population size across a region. Despite the similarity in timing of bottleneck, however, the estimates of the overall population size of these clusters differed. For example, the historic population size of the Iranian Jewish cluster was estimated to be less than 10,000 for the last 30 generations. This extremely small size could be a consequence of the historic persecution of Jews, a hypothesis in line with our estimate and other published estimates [75], [76] of the small historic population size for Ashkenazi Jews. However, population size estimates may also be biased by the selective migration of clusters to Los Angeles. Forces such as these, especially those that would affect historic mate choice, could create ascertainment biases in the patterns of IBD sharing observed here.

#### 3.4.2 Between cluster IBD

Patterns of IBD sharing between clusters can further reveal modern and historical relationships. These might be important to consider when analyzing health inequities as related groups might have similar environmental and genetic exposures. To explore this, we first computed pairwise Hudson’s F_ST_ in the largest IBD clusters (Fig. 6d). Consistent with other reports [77], we found that F_ST_ is large between IBD clusters with differential continental origins, indicating greater differentiation. In particular, the West African, African American, and Afro-Caribbean IBD clusters had the largest F_ST_ with other clusters. F_ST_ also revealed the complex sharing patterns within continental IBD clusters. While there was low differentiation between the Iranian Non-Jewish and Iranian Jewish IBD clusters (F_ST_=0.0055), the Iranian Non-Jewish cluster exhibited a smaller F_ST_ with the Armenian cluster (F_ST_ =0.0015), Lebanese Christian cluster (F_ST_ =0.0030), and Pakistani cluster (F_ST_ = 0.0038). It is important to note, however, that the F_ST_ estimates used here are from genotyping data and do not necessarily capture the effect of rare variants [77], which are important for understanding fine-scale relationships.

To further characterize the patterns of relatedness in ATLAS, we created a network representation of IBD sharing. The nodes of the network were each IBD cluster, and the edges were the median IBD shared between clusters. This network was then visualized using 1000 iterations of the Fruchterman-Reingold force-directed algorithm [78]. From this representation, we found that IBD relationships of clusters in LA recapitulated well-known relationships (Fig. 6e). We observed that geography affected cluster relationships. For example, clusters with Middle Eastern ancestry were close, with the Pakistani cluster acting as a bridge between them and the South Asian IBD clusters. The Iranian Jewish cluster and Iranian Non-Jewish cluster were also depicted as close, as well as the clusters of Ashkenazi Jews and European Non-Jews. We also observed some unexpected relationships. The Central/South American IBD cluster both shared more IBD on average with Ashkenazi Jews (mean=0.243 cM, 95% CI:[0.243, 0.244]) than European Non-Jews (mean=0.0372 cM, 95% CI: [0.0371, 0.0373). A similar trend was observed for the Puerto Rican cluster and the Ashkenazi Jewish cluster (mean=0.212 cM, 95% CI: [0.212, 0.215]) vs the European Non-Jewish cluster (mean=0.495 cM, 95% CI: [0.0489, 0.0501]). Other reports have found a contribution of Jewish ancestry to Latin American clusters [79].

### 3.5 Website

To facilitate sharing of all the associations for the major IBD clusters identified in ATLAS, we developed a website, www.ibd.la. This website has several pages that allow open-access exploration of all associations between the clusters. The pages include phecode associations for both outpatient and emergency room diagnoses, an interactive map of Los Angeles to visualize primary care offices most utilized by the clusters discussed here, and visualizations of IBD shared between clusters. Summary statistics can also be downloaded.

## 4 Discussion

To reduce health disparities and ensure that precision medicine initiatives are applicable to all people, it is important to understand the diverse determinants of health. In this study, we analyzed the healthcare utilization of clusters of individuals who share genetic ancestry. This allowed for a more fine-scale analysis into health outcomes than using race and ethnicity information typically available in the EHR. Our approach thus provides a complementary lens for identifying potential inequities in health within Los Angeles.

There are many considerations when interpreting the results of this study. It is essential to note that race, ethnicity, and religion are social constructs and are not determined by genetics, although they may be correlated [21], [24], [80], [81]. Despite this, however, it is simultaneously true that the social constructions of race, ethnicity, and religion affect healthcare in the United States [82]–[86].

To study diverse health outcomes in the context of EHRs, we defined and named IBD clusters. However, defining a population or cluster is not straightforward [19], [87]. We followed recent work and chose a genetic similarity criterion in an unsupervised machine learning algorithm to define an IBD cluster [13], but any number of criteria or algorithms could have been used. Likewise, many different names could have been chosen for a cluster, or numeric values could have been assigned. The name of the IBD cluster also does not reflect the identity of individuals within that cluster, nor do individuals within the clusters defined here necessarily have a shared sense of identity, as with the traditional sociological definition of a community [88].

We were limited to labeling IBD clusters based on demographic information contained in the EHR and reference data, which is imperfect and does not capture the full complexity of cluster identity. Historical events might also affect the representativeness of the clusters. An example is that a major wave of Armenian immigrants in Los Angeles originated primarily from Western Armenia, because of the Armenian genocide and fall of the Ottoman Empire [89]. For this reason and many others, the Armenian IBD cluster we discussed here might not be representative of Armenians outside of Los Angeles. We are not able to characterize to what extent historical events bias the results for all IBD clusters, both in terms of the accuracy of the labels we used, and in terms of the generalizability of our results beyond Los Angeles.

Furthermore, although we presented many IBD clusters, the clusters are not equivalent. Some IBD clusters identified here were tightly related in IBD network space. Others had more diffuse patterns of connection. In some cases, IBD clusters that were less well connected could represent an opportunity for further analysis. For example, China contains large genetic, linguistic, and ethnic diversity [90], yet in this study, we examined the Chinese cluster as a single unit. Future work with more data could focus on additional fine-scale clusters contained within the IBD clusters analyzed here.

There are also limitations to our healthcare utilization results. Foremost, the reported associations are strictly correlative. Although we used genetics to identify clusters, genetics is not the cause of the associations. Instead, as demonstrated by the changes in diagnoses over time, there are likely complex external processes affecting why a cluster receives a phecode or visits a specific doctor’s office.

Another limitation is that individuals who comprise ATLAS are not random. Individuals who come to a hospital are usually sick. Additionally, groups who live farther away may only be showing up to UCLA Health, which is a nationally recognized hospital [53], because they are seeking specialty care due to a more severe condition. This is compounded by the fact that the main UCLA Health facilities are in west Los Angeles, which includes some of the wealthiest neighborhoods in Los Angeles County by median income, including Beverly Hills and Bel-Air [91]. Thus, poorer clusters might be traveling farther to access care and have greater health needs motivating the longer trip. These factors would directly bias the associations that we observed. Additionally, the observation that some groups have greater travel time to reach specialty care could be the basis of future studies on health equity.

Insurance coverage dramatically influences access to care in the United States [92] and coverage varies by race and ethnicity [93]. As a result, some IBD clusters may have access to more comprehensive insurance and thus receive more specialized diagnoses, or more diagnoses in general. Unfortunately, detailed insurance information is not available in this data. Other socioeconomic factors, such as age, education, and household income are also associated with when and if patients receive diagnoses [94]–[96]. Therefore, differences in cluster demographics likely affect these results. These differences may also be exacerbated by biases that exist on the part of health practitioners, as implicit biases have been shown to systematically affect care [66], [97]–[100].

Lastly, our results may be limited by the nature of the data used, namely, genotyping data. There are other ways we could have defined clusters, such as designing self-identification surveys. For looking at pathogenic alleles, we were restricted in the number of sites we could consider, and potentially biased by the fact that genotyping chips are generally ascertained for sites that are common and shared between populations [101]. This could also bias our IBD detection as the density of genotyped sites or overall quality could affect the IBD calling [12]. Furthermore, statistical phasing with publicly available genetic maps may have reduced accuracy in populations that are less related to those contained in the reference panel [102].

There are many areas of future directions for this work. Firstly, additional work should be done to confirm these results and design interventions that address potential inequities. IBD clusters may also be useful in controlling for population stratification in other studies examining the social determinants of health. Environmental factors were not explored in this study but could be an avenue for future research as they impact health, often inequitably. For example, air pollution poses a substantial risk to human health [103]. Air pollution levels vary substantially across the United States and toxic levels disproportionately affect communities of color [104]–[106]. Thus, understanding environmental factors could provide greater context to the health outcomes examined here.

Lastly, while every participant is placed into a cluster, this approach may be limited for individuals with multiple ancestries. For example, someone with one parent who has predominantly Iranian Jewish ancestry and one parent with predominantly Japanese ancestry would be assigned to either the Japanese or Iranian Jewish cluster in our current approach, which would not capture the full complexity of their true ancestry. These individuals represent a crucial group in the advancement of precision medicine [107] and future work could focus on opportunities to study the health of these admixed individuals.

Overall, we identified and characterized the health profiles of Los Angeles IBD clusters, many of whom, to our knowledge, have not been studied in the context of a biobank before. This represents an advance toward equitable health research and, along with our website, can empower future studies on health outcomes in Los Angeles.

## 5 Methods

### 5.1 Patients and Recruitment

### 5.2 EHR Data

Each patient’s genotype data was tied to Electronic Health Records (EHR) using a de-identified ID. Patient EHR was pulled for 2016-2020 and included visit information, diagnosis information, and demographics. For the normal outpatient data, we restricted to visits that were labeled as scheduled appointments and that did not have a code associated with an inpatient, ICU, or trauma stay. Emergency room data was any visit that happened within an emergency room department. Diagnoses assigned in emergency rooms were restricted to the primary reason for the visit. Each visit contained information on patient weight, height, and BMI measured at the visit. We calculated the median BMI for a patient across all encounters and used this as the BMI for that patient in our association testing.

Demographic information was restricted to race/ethnicity, preferred religion, preferred language, sex, and birth date. Sex was indicated as binary. To calculate patient age, we calculated the patient age at the time of each visit and took the maximum age overall for each patient. For EHR reported race/ethnicity, patients were designated (by themselves or a healthcare staff member) as “White,” “Black”, “Asian”, “Native American”, or “Pacific Islander.” Asian patients could be further designated as Chinese, Japanese, Korean, Thai, Filipino, Vietnamese, Taiwanese, Pakistani, Indian, or Indonesian, although not all Asian patients had one of these identifiers. Hispanic patients were designated as “Hispanic”, which was further subdivided into several other sub-identifiers, such as “Spanish origin”, “Chicano/a” or “Cuban”. For visualization, we considered the main race/ethnic categories and not the sub-designations. There were numerous preferred languages and religions. For simplicity, we examined the languages that had more than 5 individuals who indicated that they preferred that language. Furthermore, preferred religion was restricted to consider major religions: Christianity, Islam, Judaism, Hinduism, Sikhism, and Buddhism. Christianity was further subdivided into Protestant or Catholic. Other religions were condensed into an “Other Religion” category.

### 5.3 Pre-processing and quality control

Genotyping for ATLAS was performed on a custom genotyping chip, with sites from the global screening array. Data was mapped to hg38 and all SNPs were mapped to the 147 build of dbSNP [108]. All preprocessing and quality control steps were performed using PLINK 1.9 [109], [110], and bcftools v1.9 [111].

For ATLAS samples, we removed any individuals whose genotyped sex mismatched their EHR reported sex. We did this by using the PLINK --update-sex command to update the PLINK fam files to contain the EHR sex and the PLINK --check-sex to identify samples with discrepancies between the estimated genotype sex and EHR sex.

ATLAS data was merged with genotyping data from the 1000 Genome Project (1000GP) [26], the Simons Genome Diversity Project (SGDP) [27], and the Human Genome Diversity Project (HGDP) [28]. All reference data were converted to hg38 for merging using CrossMap [112]. Samples that overlapped between the different projects were removed using PLINK --keep. Rsids were harmonized across projects using bcftools annotate. Data were then standardized using bcftools norm and a hg38 genome reference. After merging, sites or individuals with more than 1% missing were removed using plink --mind and --geno. For IBD analysis, only SNPs with MAF > 5% were kept.

Before IBD calling, data was statistically phased using Shapeit4 [113] using default parameters and the hg38 map files distributed with the software. To speed up computation, one chromosome was phased at a time.

### 5.4 IBD calling and processing

For IBD calling, the genotype data were converted from PLINK bed files into PLINK ped/map files using a custom Python script that preserves phasing. Centimorgan information for the map files was pulled from the same genetic maps used in Shapeit4.

IBD was called using iLASH [29] with the following parameters: slice_size 350, step_size 350, perm_count 20, shingle_size 15, shingle_overlap 0, bucket_count 5, max_thread 20, match_threshold 0.99, interest_threshold 0.70, min_length 2.9, auto_slice 1, slice_length 2.9, cm_overlap 1, minhash_threshold 55. IBD was called for one chromosome at a time.

After IBD was called, we removed outliers as in Belbin et al [11]. Firstly, any IBD segments overlapping centromeres or telomeres were removed. IBD tracts intersecting the HLA region were also removed. To find other regions of the genome that may have erroneously high IBD, we calculated the total amount of IBD contained at each SNP in our input file by summing all segments that overlapped that SNP. SNPs that had total IBD greater or less than 3 standard deviations from the genome-wide mean were removed. In total, 6696 were removed.

For downstream analysis, IBD segment lengths were summed between individuals, meaning that for a given pair of individuals, all the IBD segments that they shared across all chromosomes were added together to create one summary number.

We removed pairs of individuals who were immediate family members using two methods. Firstly, we used the KING relatedness inference software [114] to find any pairs of individuals who were closer than second-degree relatives, using the parameters --kinship --degree 2. King was run on all SNPs with MAF > 0.05 and after linkage pruning, using PLINK and --indep-pairwise 50 10 0.1. As KING may underestimate the relatedness of individuals, especially in the case of individuals with high levels of autozygosity [29], we also filtered pairs based on the total amount of IBD shared. Using empirical data reported to DNA Painter [115], we determined a conservative threshold of second-degree relatedness was a threshold of 1000cM. We removed any pairs with IBD higher than this threshold.

### 5.5 Cluster identification and annotation

For cluster detection, we used the Python package NetworkX [116]. We created an undirected graph representation of our IBD matches, where each node was an individual and an edge between individuals was weighted by the total amount of IBD matches shared between the two people.

Louvain clustering [30], implemented in NetworkX, was used iteratively to detect fine-scale populations. It was first run to detect a primary set of clusters. Each cluster was then subject to Louvain clustering again, and these subclusters were clustered once more, for a total of three runs of Louvain clustering.

After generating clusters with the Louvain algorithms, the clusters were merged using F_ST_, as in Dai et al [13]. We used the implementation of Hudson’s F_ST_ from PLINK 2.0. It was run on all pairs of clusters from the third level of the Louvain clustering and clusters that had F_ST_ < 0.001 were merged. Since F_ST_ may perform poorly in small populations, clusters with less than 10 people were ignored [77]. This threshold was selected because it gave good separation of clusters on a subcontinental level.

Once clusters were merged, they were annotated with the demographic information from the EHR. For the individuals in the IBD cluster that did not prefer English, we calculated the proportion of individuals who preferred each of the 20 languages spoken by more than 5 biobank individuals. Similarly, after excluding individuals who did not have information on religion in the EHR, we calculated the proportion of individuals in that cluster who preferred one of the major religions indicated in our demographic EHR data. Using this data, race/ethnicity information, and the reference populations that segregated in a cluster, we holistically assigned a population name to an IBD cluster. The names were chosen to correspond to a modern country that had historical or modern immigration into Los Angeles (i.e., “Chinese”), or clusters that make up well-known demographic slices of America (i.e. European Non-Jewish). When no country of origin or demographic was obvious given the available data, we named the cluster with what distinguishing information was available (i.e., “Central/South American”).

For downstream analysis, we focused on IBD clusters that had more than 40 members to ensure large enough sample size for our EHR and genetic analyses.

### 5.6 Genetic analyses

To find the distribution of IBD in a cluster, we considered IBD segments of individuals assigned to the same cluster. We summed the IBD segments to get the total IBD shared between the pair and calculated the distribution of total IBD between members of the cluster.

For ROH, we first performed linkage pruning and MAF filtering using PLINK and the parameters --maf 0.01 --indep-pairwise 50 10 0.1. ROH calling was also performed using PLINK and the parameters -homozyg --homozyg-density 200 --homozyg-gap 500 --homozyg-kb 3000 --homozyg-snp 65 -homozyg-window-het 0 --homozyg-window-missing 3 --homozyg-window-snp 65. Detected ROH were summed within an individual. We then calculated the distribution of detected ROH of all individuals within a cluster.

IBDNe was run using the IBD haplotypes estimated using iLASH [74]. We filtered the iLASH output for each chromosome to individuals from a single cluster. The haplotypes were combined into one file for IBDNe input. IBDNe was run with default parameters and the hg38 genetic map provided on the IBDNe website.

For the heatmap of F_ST_, we calculated the pairwise Hudson’s F_ST_, as described in the Louvain clustering section. We calculated F_ST_ between the largest final clusters (after Louvain clustering and merging). Data was visualized using Python Seaborn clustermap with default parameters [117].

The network visualization between IBD clusters was developed using NetworkX. The input was a matrix where each row and columns represented one of the largest clusters, and each entry was the mean IBD shared between the two clusters. To find this mean, we found all possible pairs of individuals between the two clusters. If the pair did not have any IBD detected, we set their IBD to 0 and then calculated the mean over all possible pairs. This was to prevent biasing the mean IBD by limiting it to only pairs that had IBD detected. This square matrix was then used to create a weighted undirected graph, where the nodes were the IBD clusters, and the edges were the mean IBD between the clusters. We visualized the graph using 1000 iterations of the Fruchterman-Reingold force-directed algorithm [78].

### 5.7 EHR association analyses

Statistical testing was done using the Python StatsModel package [118]. For each phecode, we determined whether an individual has ever had been assigned that phecode in an outpatient context, making the outcome binary. Cluster status was binary and could either be a particular cluster vs all other biobank participants, or a particular cluster compared against another cluster. We tested whether binary cluster status was associated with phecode assignment using the StatsModel GLM command with the family set to binomial. We corrected for sex, age, and BMI in these analyses. Specifically the command we used was: GLM.from_formula(“phecode_status ∼ cluster_status + sex + age + bmi”, family= sm.families.Binomial(), data=model_input).

The same statistical framework was used to test for emergency room diagnoses and specialty visits, where instead of phecode assignment, the outcome was whether or not an individual had visited a doctor with a given specialty reported in the EHR. An association was considered significant after controlling for false discovery rate at 5%.

### 5.8 Zip Codes

For analyzing the zip codes associations, each office or department visited by biobank patients was associated with a primary zip code. We excluded the two main UCLA hospitals, since we were looking for differential utilization across the greater Los Angeles area. For plotting, we associated these zip codes with geographic coordinates using a shapefile obtained from the Los Angeles GeoHub [119], which was loaded in Python using the GeoPandas package [120]. Specifically, we used the EPSG:3857 coordinate system, which is widely used by map providers.

For each zip code, the StatsModel GLM command was used to estimate the association between cluster status and ever visiting a particular zip code relative to the whole biobank or opposing IBD cluster, corrected for sex, age, and BMI. GeoPandas was used to plot, where the color of the zip code either corresponded to the cluster that associated with a particular zip code, or its intensity corresponded to the log odds ratio of the association between a particular cluster visiting offices in a zip code. Basemaps were added to the plot using the Python package Contextily [121] and the Toner Lite open-source map data from Stamen Design [122]

### 5.9 Website

The website hosting the data visualization is implemented as a single-page application [123]. The application is developed in the JavaScript framework React, where each graph page is implemented as a separate component. The map plot is powered by the deck.gl library [124] developed by Mapbox, which provides maps for data overlays. The other graphs are powered by the react-plotly.js library developed by Plotly [125], which provides a React interface to create interactive plots. The application has no backend, as the data is relatively small, requires no modification or manipulation per request, and is not subject to any privacy concerns due to its approval for release. All the data is stored in static JSON files that the application directly references to generate data visualizations. The website code and underlying data are publicly available on Github with an MIT license, which will allow others to contribute to the application as well as use the code to build visualizations for their own organizations.

### 5.10 Data Visualization

Data analysis was done in Python 3.8 [126] using Jupyter Notebooks [127]. Visualization was done using Seaborn [117] and Matplotlib [128].

## Data Availability

Patient level EHR and genotyping data is protected due to patient privacy. Summary statistic data is available on www.ibd.la

## 6 Acknowledgements

We thank Vivek Kumar and Michael Broudy for their expertise with the DDR. We thank Aaron Panofsky and Anna Lewis for their helpful comments and discussion on this manuscript. We gratefully acknowledge the Institute for Precision Health, participating patients from the UCLA ATLAS Precision Health Biobank, UCLA David Geffen School of Medicine, UCLA Clinical and Translational Science Institute, and UCLA Health.

C.C. was supported by F31NS122538. C.C., N.Z., D.E., and E.P. were supported by grants from the NIH and DoD: R01CA227237, R01ES029929, R01MH122688, U01HG009080, R01HL155024, R01HL151152. R01GM142112, R01HG006399. N.Z., E.K., C.G., V.A., G.B. were supported by R01HG011345. A.C. was supported by T32HG002536 and DGE-1829071. V.A. was supported by DP5OD024579.

## 7 Software

Code for IBD calling and clustering is available at https://github.com/christacaggiano/IBD. Code for the website is available at https://github.com/misingnoglic/ibd.la.

## 8 Supplementary Figures

**Figure S1:**
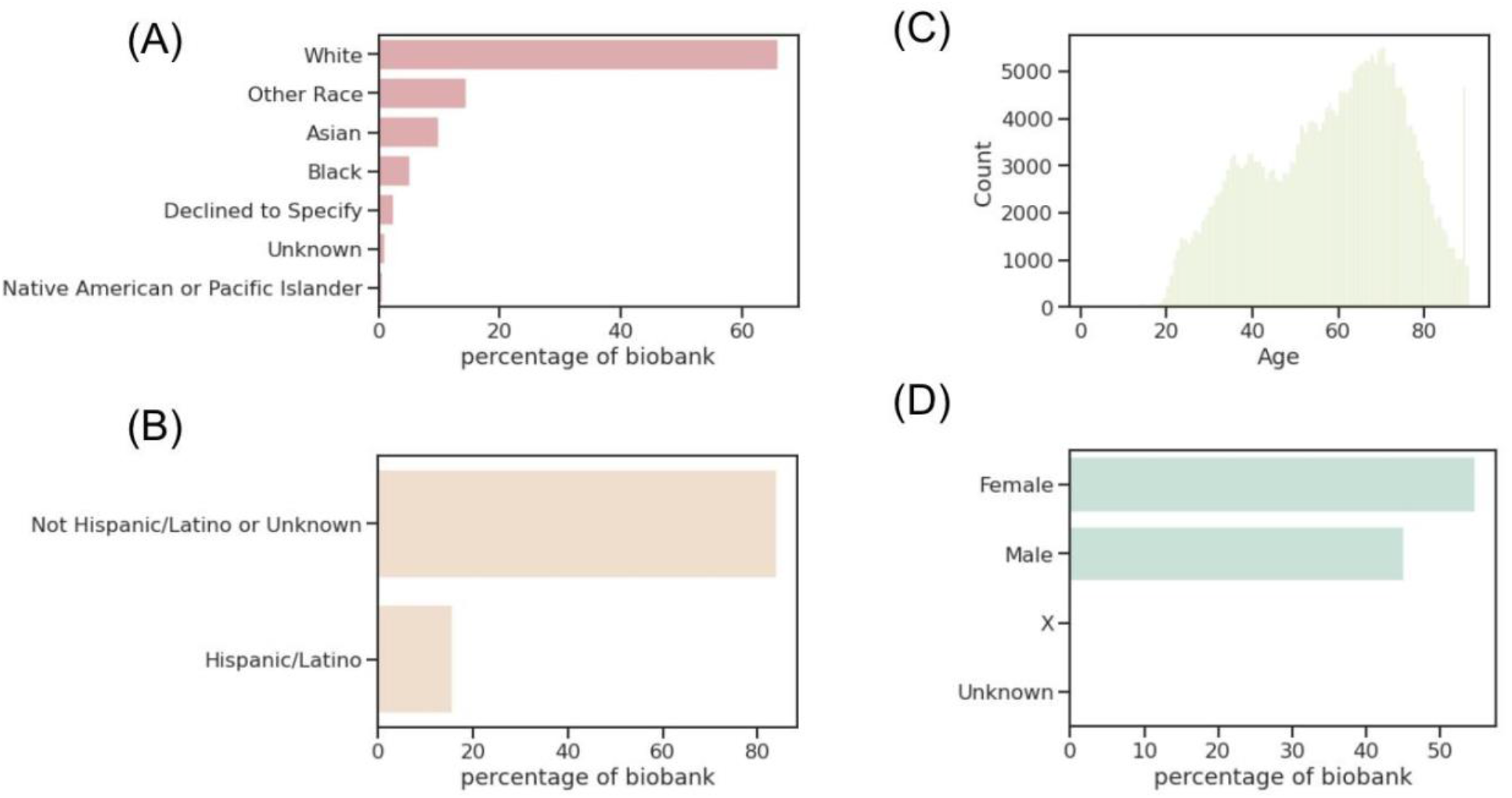
ATLAS demographics. (A) The percentage of ATLAS that is recorded as each race, (B) the percentage of ATLAS recorded as Hispanic, (C) the distribution of patient age in ATLAS, and (D) the percentage of ATLAS recorded as each sex.

**Figure S2:**
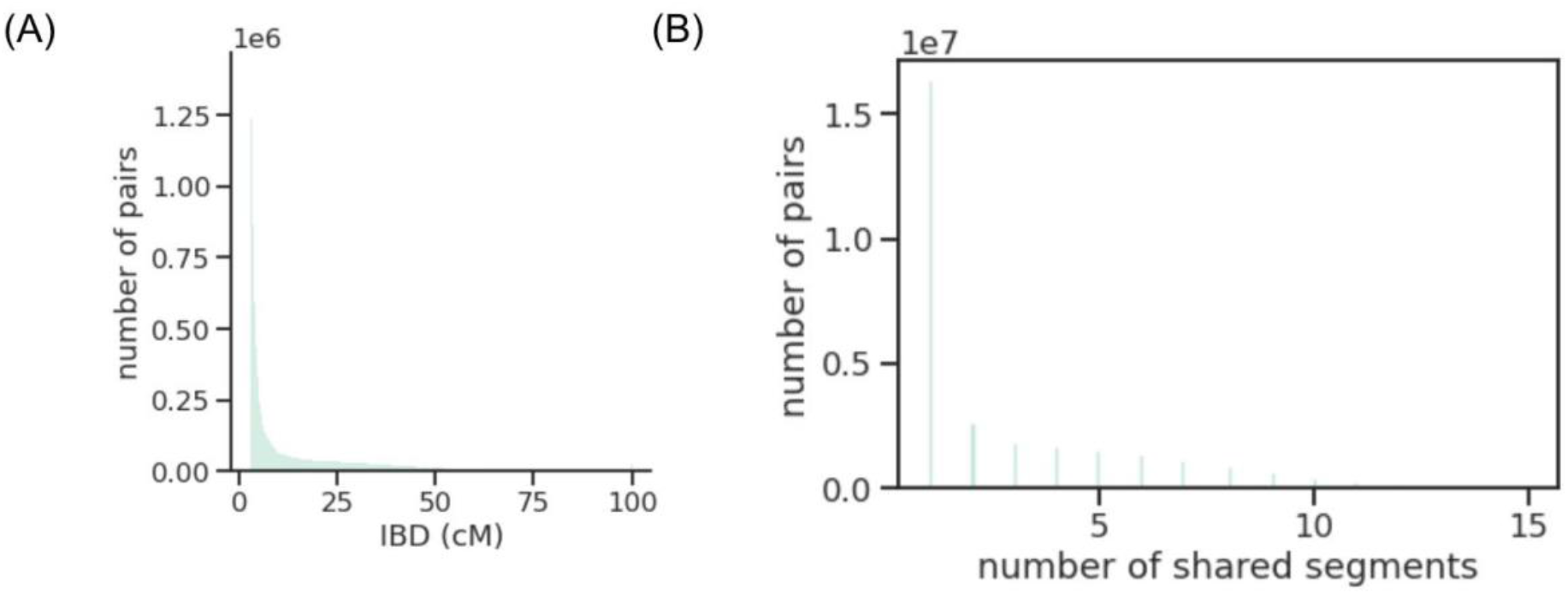
Overall IBD length and segment distribution. (A) The distribution of total pairwise IBD estimated for all biobank and reference participants and (B) The distribution of the total number of IBD Segments shared between pairs.

**Figure S3:**
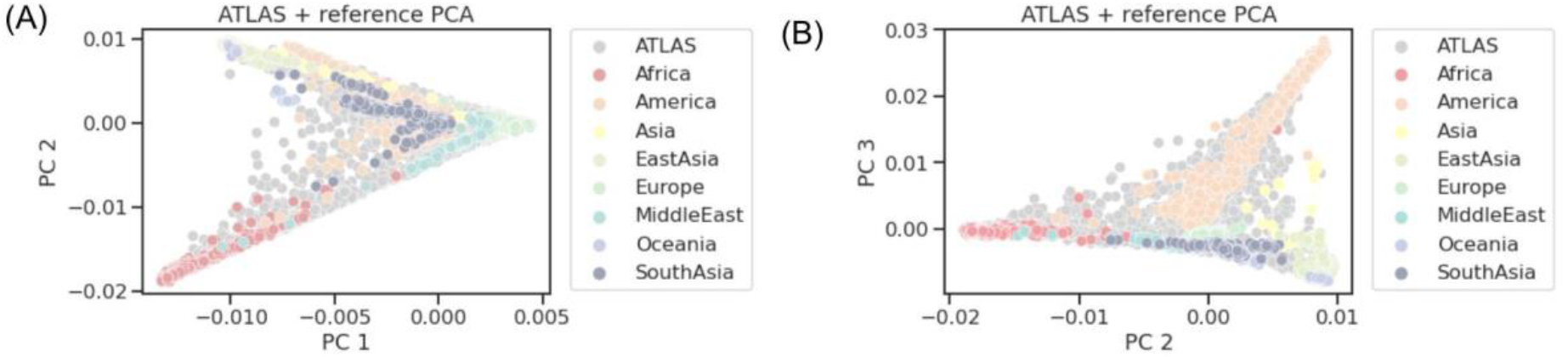
Principal component analysis of ATLAS and reference data. (A) Principal component (PC) 1 vs PC2 and (B) PC2 versus PC3.

**Figure S4:**
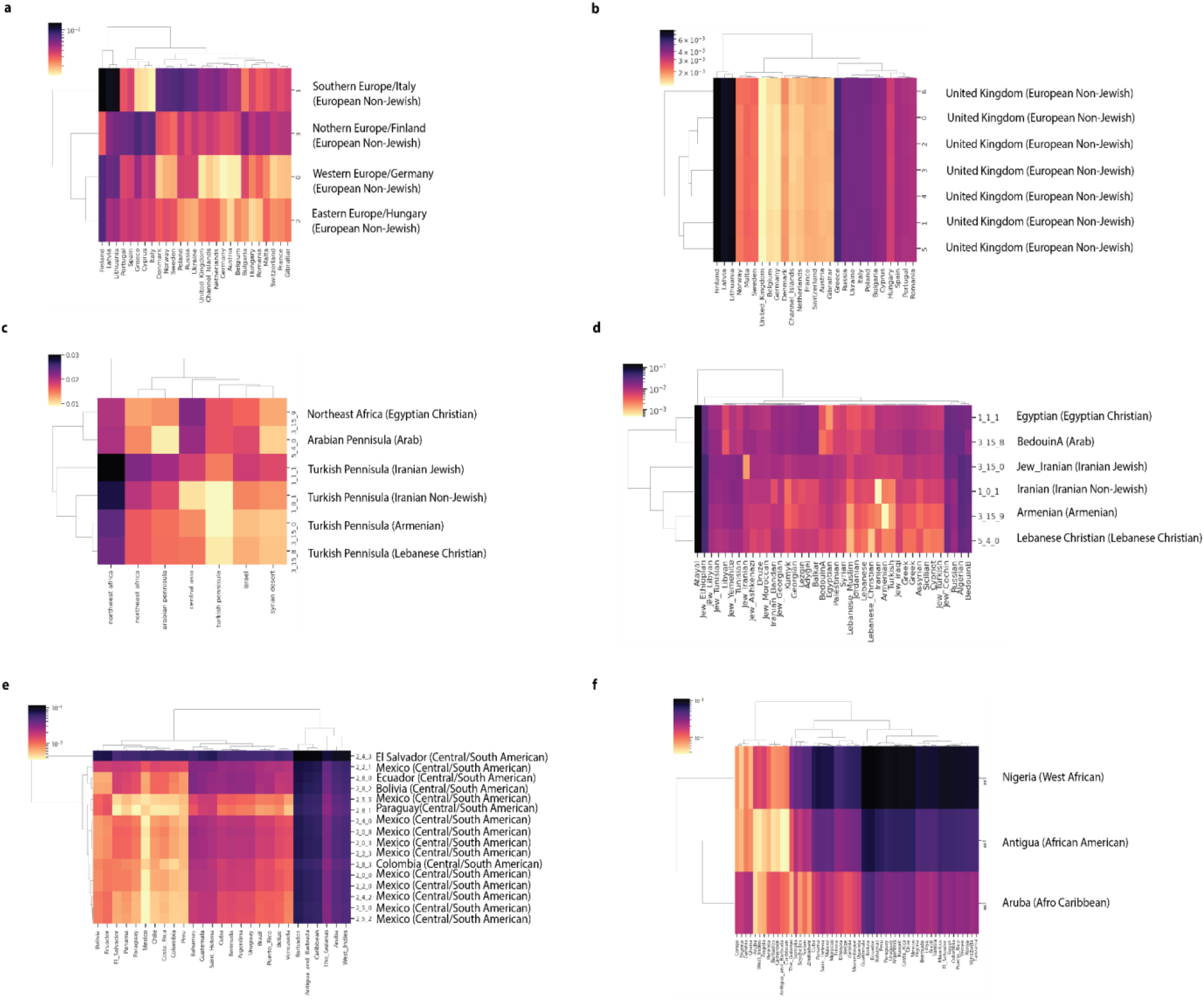
F_ST_ between IBD clusters and external reference data. (a) F_ST_ between one set of subclusters (subclusters UCLA_3_7_*) that made up the European Non-Jewish IBD cluster and samples from the UKBB [129] who were born outside the United Kingdom, combined with a random sample of 100 individuals born in the United Kingdom. A second set of European subclusters (subclusters UCLA_3_8_*) are shown in (b). (c) F_ST_ between the Greater Middle East Variome [130] populations and UCLA IBD clusters with Middle Eastern or Central Asian ancestry and (d) F_ST_ between modern day Middle Eastern populations [131] and UCLA IBD clusters with Middle Eastern/Central Asian ancestry. (e) F_ST_ between UKBB participants born in the Americas and subclusters that made up the Central/South American cluster. (f) F_ST_ between UKBB participants born in Africa or the Americas and the three Black/African American clusters. For all plots, the country with the smallest F_ST_ to the ATLAS cluster is labelled. The ATLAS cluster name the subcluster belongs to is indicated in parentheses. The brighter the color, the smaller the F_ST_ value, suggesting less differentiation between the two groups.

**Figure S5:**
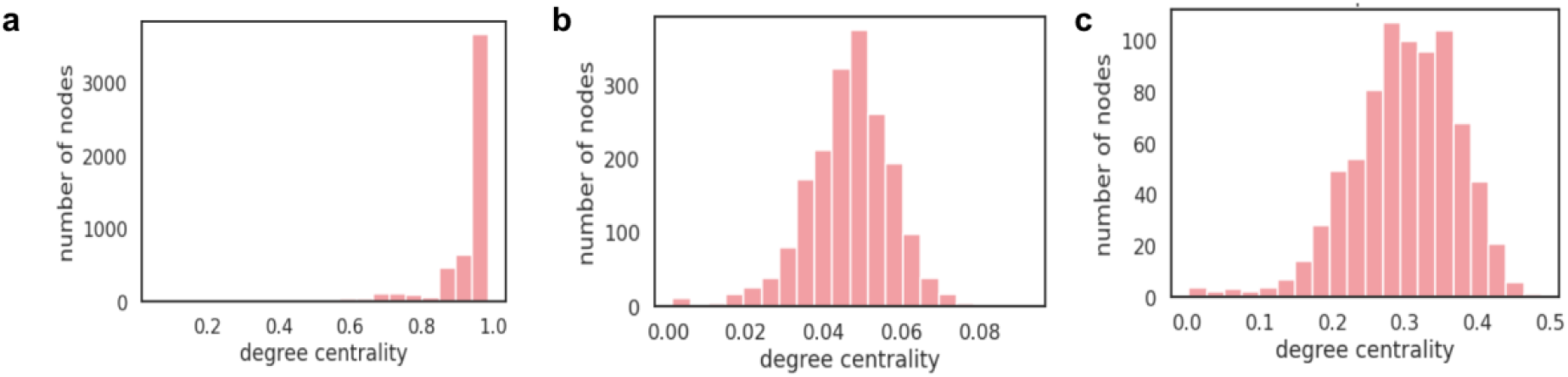
Degree centrality of clusters. The degree centrality distribution (node degree divided by the max possible degree in the cluster) of selected clusters from the final round of Louvain clustering. (A) is a cluster where nearly every individual in the cluster is connected to every other member of the cluster. (B) is an example of a cluster where individuals share some connections, but on average are less connected to each other, and (C) is an example where individuals are moderately connected to each other.

**Figure S6:**
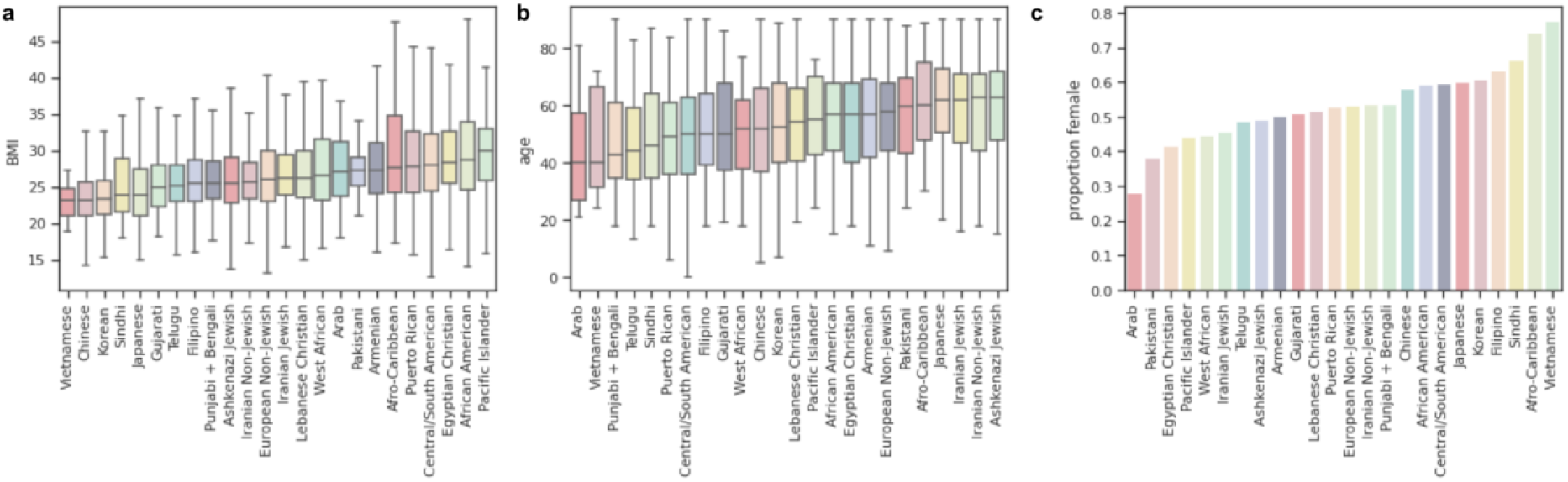
Demographics of IBD clusters. For each of the largest IBD clusters, the (a) distribution of median patient BMI of participants in the cluster, (b) the distribution of max patient age of participants in the cluster, and (c) the proportion of the cluster that is female based on EHR demographic records.

**Figure S7:**
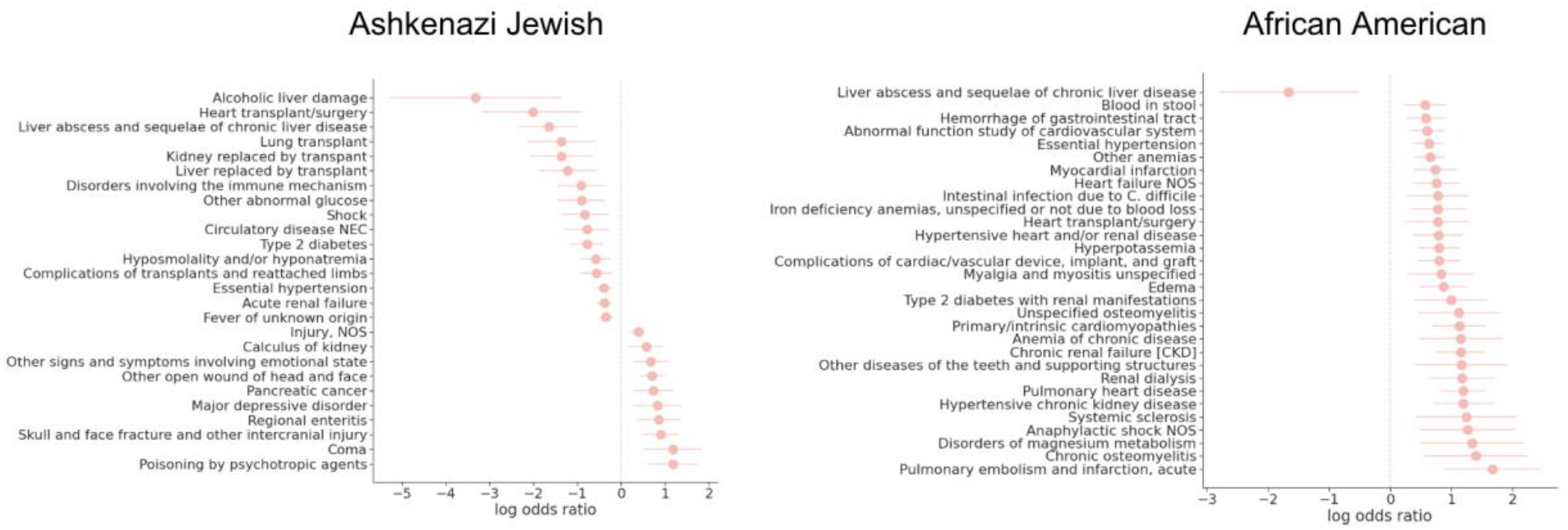
Phecodes associated with Armenian cluster status in different contexts. The log odds ratio of whether a given phecode assignment is associated with membership in the (A) Ashkenazi Jewish IBD cluster and the (B) African American IBD cluster versus membership in the remaining biobank, in emergency room settings. A log odds ratio greater than zero means that the phecode is more likely to be given to patients of that cluster. We tested only phecodes with at least 30 patients per code and corrected for age, sex, and BMI in these analyses. Phecodes significant at FDR 5% are shown and if there are more than 30 significant associations, we plot only the top 40 with the largest absolute log odds ratio.

**Figure S8:**
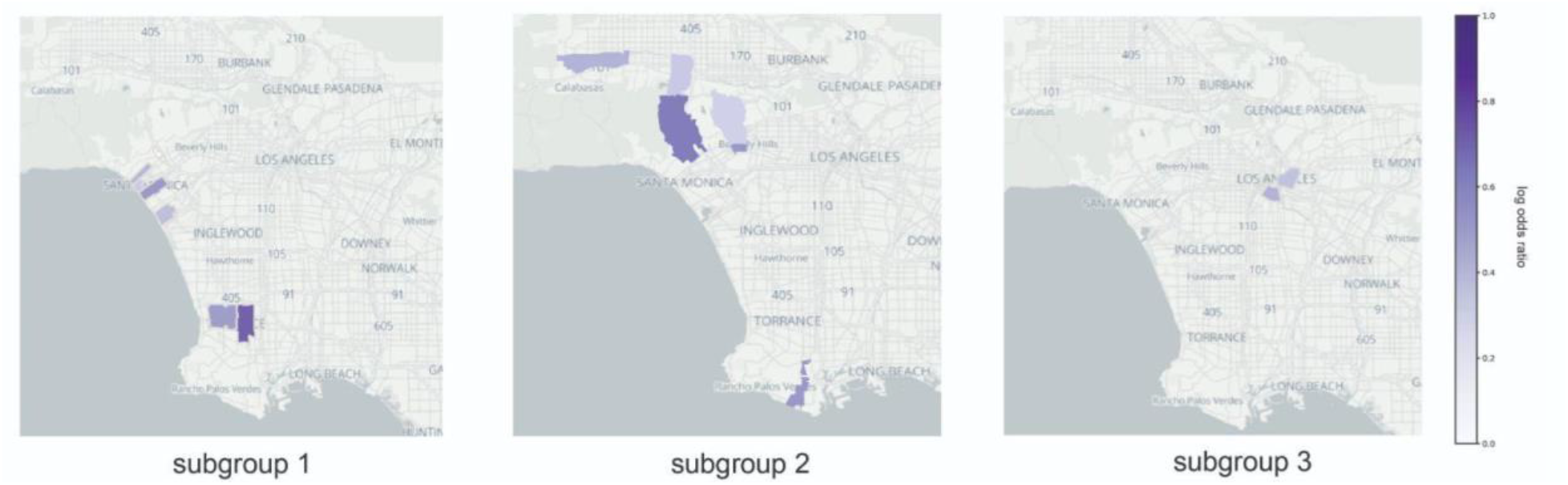
Central/South American IBD cluster subgroup office utilization. For three FST subgroups of the Central/South American IBD cluster, the offices that they are most likely to visit. The darker the color on the graph, the more likely that cluster is to visit that office relative to other individuals in this cluster at UCLA.

**Figure S9:**
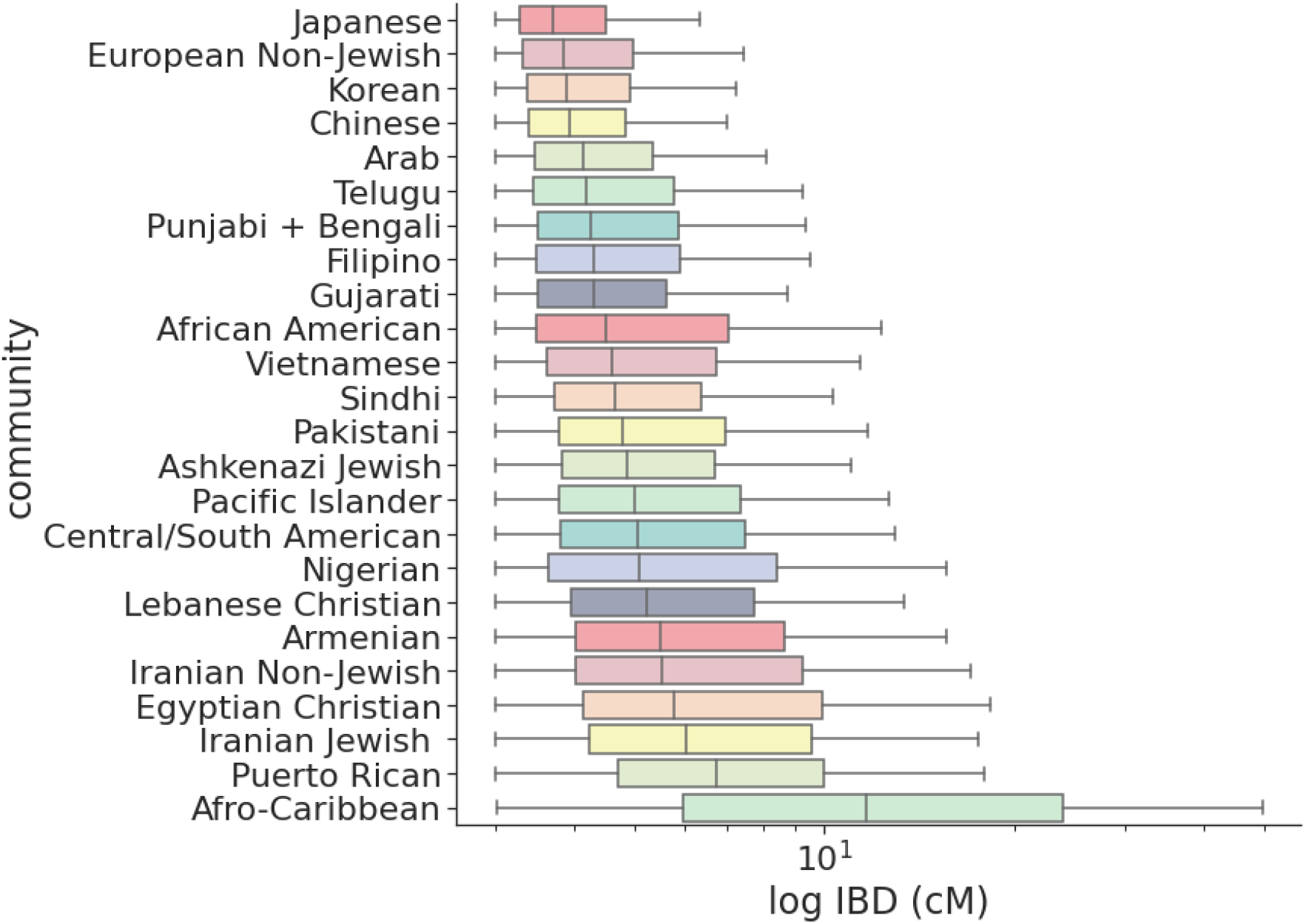
Distribution of IBD per cluster. The log-scale distribution of IBD segment lengths in each of the largest IBD clusters.

## 9 Supplementary Tables

**Table S1:**
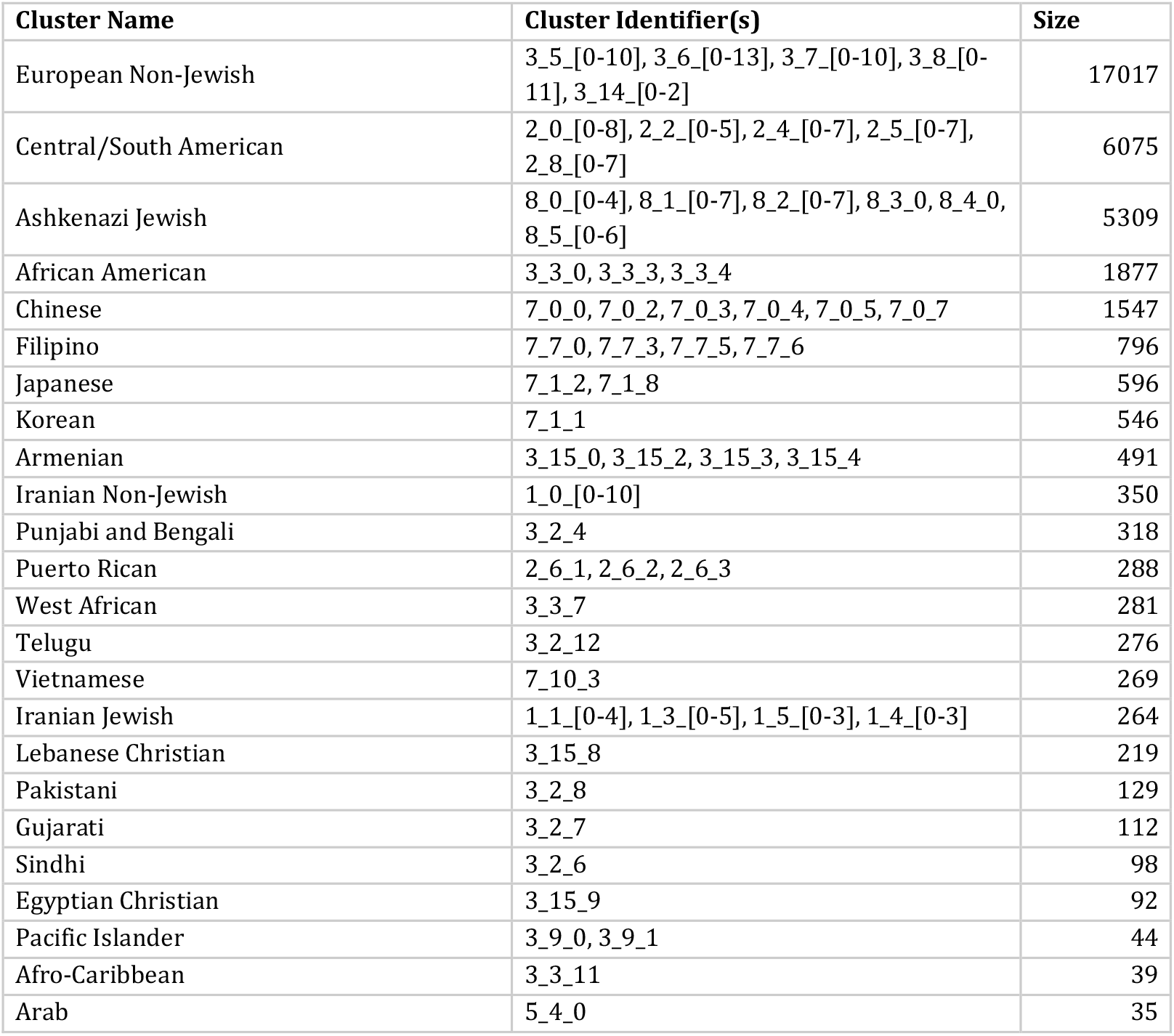
Largest ATLAS IBD clusters. For the 24 largest ATLAS IBD clusters, the identifier from the Louvain algorithm, the name we assigned the IBD cluster, and the total size of the IBD cluster.

| Cluster Name | Cluster Identifier(s) | Size |
| --- | --- | --- |
| European Non-Jewish | 3_5_[0-10], 3_6_[0-13], 3_7_[0-10], 3_8_[0-11], 3_14_[0-2] | 17017 |
| Central/South American | 2_0_[0-8], 2_2_[0-5], 2_4_[0-7], 2_5_[0-7], 2_8_[0-7] | 6075 |
| Ashkenazi Jewish | 8_0_[0-4], 8_1_[0-7], 8_2_[0-7], 8_3_0, 8_4_0, 8_5_[0-6] | 5309 |
| African American | 3_3_0, 3_3_3, 3_3_4 | 1877 |
| Chinese | 7_0_0, 7_0_2, 7_0_3, 7_0_4, 7_0_5, 7_0_7 | 1547 |
| Filipino | 7_7_0, 7_7_3, 7_7_5, 7_7_6 | 796 |
| Japanese | 7_1_2, 7_1_8 | 596 |
| Korean | 7_1_1 | 546 |
| Armenian | 3_15_0, 3_15_2, 3_15_3, 3_15_4 | 491 |
| Iranian Non-Jewish | 1_0_[0-10] | 350 |
| Punjabi and Bengali | 3_2_4 | 318 |
| Puerto Rican | 2_6_1, 2_6_2, 2_6_3 | 288 |
| West African | 3_3_7 | 281 |
| Telugu | 3_2_12 | 276 |
| Vietnamese | 7_10_3 | 269 |
| Iranian Jewish | 1_1_[0-4], 1_3_[0-5], 1_5_[0-3], 1_4_[0-3] | 264 |
| Lebanese Christian | 3_15_8 | 219 |
| Pakistani | 3_2_8 | 129 |
| Gujarati | 3_2_7 | 112 |
| Sindhi | 3_2_6 | 98 |
| Egyptian Christian | 3_15_9 | 92 |
| Pacific Islander | 3_9_0, 3_9_1 | 44 |
| Afro-Caribbean | 3_3_11 | 39 |
| Arab | 5_4_0 | 35 |

**Table S2:** Log odds ratio of visiting an emergency room. Using a logistic regression model controlling for age, sex, and BMI, the p-value and log odds ratio of membership in an IBD cluster being associated with visits to an emergency room, relative to the remaining biobank participants.

| IBD cluster | p-value | log odds ratio |
| --- | --- | --- |
| Korean | 9.82E-01 | -0.0023+/-0.1945 |
| Filipino | 9.32E-01 | 0.0069+/-0.1534 |
| Telugu | 9.25E-01 | -0.017+/-0.373 |
| Puerto Rican | 9.17E-01 | -0.0178+/-0.352 |
| Lebanese Christian | 8.45E-01 | 0.0296+/-0.267 |
| Sindhi | 8.38E-01 | -0.0517+/-0.5468 |
| Pacific Islander | 8.12E-01 | 0.0837+/-0.6076 |
| Gujarati I | 7.75E-01 | 0.0655+/-0.3838 |
| Japanese | 7.42E-01 | 0.0336+/-0.1666 |
| Ashkenazi Jewish | 5.29E-01 | 0.0209+/-0.0442 |
| Punjabi + Bengali | 4.71E-01 | 0.1181+/-0.2033 |
| Pakistani | 4.69E-01 | 0.2658+/-0.454 |
| Gujarati II | 3.62E-01 | -0.4216+/-1.3273 |
| Vietnamese | 3.51E-01 | -0.9986+/-3.0984 |
| West African | 2.36E-01 | 0.3663+/-0.2394 |
| Colombian | 1.40E-01 | -0.9225+/-2.1475 |
| Arab | 9.32E-02 | 0.6147+/-0.1029 |
| Iranian Non-Jewish | 4.45E-02 | 0.2508+/-0.0062 |
| Afro-Caribbean | 1.44E-02 | 0.857+/-0.1708 |
| Armenian | 1.21E-02 | 0.2445+/-0.0534 |
| Egyptian Christian | 9.97E-05 | 0.8507+/-0.4222 |
| Chinese | 1.97E-05 | -0.2919+/-0.426 |
| Iranian Jewish | 1.33E-06 | 0.5753+/-0.3421 |
| African American | 8.72E-30 | 0.5958+/-0.4928 |
| Central/South American | 4.74E-77 | 0.5957+/-0.5329 |
| European Non-Jewish | 4.22E-89 | -0.4902+/-0.5382 |

**Table S3:** IBD and ROH within clusters. For the 24 largest ATLAS IBD clusters, the mean total pairwise IBD detected between individuals in the cluster and the mean ROH detected within individuals of the cluster.

| Cluster | IBD (cM) | IBD standard error (cM) | ROH (MB) | ROH standard error (MB) |
| --- | --- | --- | --- | --- |
| Arab | 11.04 | 0.79 | 88.13 | 17.61 |
| Armenian | 10.63 | 0.20 | 37.64 | 3.39 |
| Ashkenazi Jewish | 26.08 | 0.00 | 12.08 | 0.29 |
| African American | 6.50 | 0.03 | 25.22 | 2.67 |
| Afro-Caribbean | 50.66 | 7.80 | 35.05 | 14.42 |
| West African | 10.07 | 0.40 | 33.89 | 3.67 |
| Chinese | 4.79 | 0.01 | 17.88 | 1.68 |
| Egyptian Christian | 11.81 | 1.54 | 62.44 | 13.07 |
| European Non-Jewish | 4.84 | 0.00 | 11.44 | 0.57 |
| Filipino | 7.23 | 0.03 | 19.75 | 1.74 |
| Gujarati | 10.03 | 0.71 | 42.48 | 4.34 |
| Iranian Jewish | 57.43 | 0.32 | 53.15 | 3.58 |
| Iranian Non-Jewish | 15.70 | 0.59 | 54.25 | 4.71 |
| Japanese | 4.81 | 0.02 | 25.29 | 2.61 |
| Korean | 4.90 | 0.05 | 27.63 | 3.27 |
| Lebanese Christian | 10.95 | 0.33 | 46.07 | 6.78 |
| Central/South American | 7.78 | 0.01 | 17.05 | 0.71 |
| Pacific Islander | 25.74 | 1.15 | 45.60 | 5.29 |
| Pakistani | 17.11 | 0.71 | 66.58 | 5.59 |
| Puerto Rican | 23.06 | 0.11 | 13.27 | 1.31 |
| Punjabi + Bengali | 8.27 | 0.42 | 42.77 | 3.96 |
| Sindhi | 17.66 | 0.30 | 41.77 | 4.38 |
| Telugu | 8.34 | 0.31 | 41.67 | 3.74 |
| Vietnamese | 10.90 | 0.54 | 38.12 | 4.02 |

## Notes

### Competing Interest Statement

The authors have declared no competing interest.

### Author Declarations

Patient Recruitment and Sample Collection for Precision Health Activities at UCLA is an approved study by the UCLA Institutional Review Board (UCLA IRB). IRB#17-001013

